# Alcohol consumption during pregnancy dysregulates maternofetal angiogenic and inflammatory factors with sex specificities

**DOI:** 10.64898/2026.07.15.26357094

**Authors:** Camille Sautreuil, Céline Lesueur, Gaëlle Pinto Cardoso, Henri Bruel, Valérie Biran, Jean-Baptiste Muller, Anne-Laure Duigou, Valérie Datin-Dorriere, Éric Verspick, Florent Marguet, Annie Laquerriere, Pierre Gressens, Bruno José Gonzalez, Stéphane Marret

**Affiliations:** Univ Rouen Normandie, Normandie Univ, INSERM U1245, Team Epigenetics and Pathophysiology of Neurodevelopmental Disorders, F-76183 Rouen, France; Univ Rouen Normandie and CHU Rouen, Department of Neonatal Pediatrics and Intensive Care, Rouen University Hospital, F-76000 Rouen, France; Univ Rouen Normandie and CHU Rouen, Department of Metabolic Biochemistry, Rouen University Hospital, F-76000 Rouen, France; Department of Neonatology, Le Havre Hospital F-76600 Le Havre, France; Department of Neonatal Pediatrics and Intensive Care, Robert Debré Hospital, AP-HP, Université Paris-Cité, INSERM U1141, F-75019 Paris, France; Université Paris Cité, Inserm, NeuroDiderot, F-75019 Paris, France; Department of Pediatrics, CHRU Morvan Hospital, F-29200 Brest, France; Department of Neonatology, Caen University Hospital, F-14033 Caen, France; Univ Rouen Normandie and CHU Rouen, Department of Obstetrics, Rouen University Hospital, F-76000 Rouen, France; Department of Pathology, Rouen University Hospital, F-76000 Rouen, France

**Author notes:** **Address correspondence to:** Bruno J Gonzalez, Inserm U1245, Faculty of Medicine, 76183 Rouen, France and Stéphane Marret, Department of Neonatal Pediatrics and Intensive Care, Rouen University Hospital, 76000 Rouen, France. Equally contributed to this work (Bruno José Gonzalez, Stéphane Marret).

**Keywords:** FASD, prenatal alcohol-exposure, neuroplacentology, angiogenesis, endothelial inflammation

## Abstract

Prenatal alcohol exposure (PAE) is a major cause of neurodevelopmental disorders, yet most children are diagnosed late or misdiagnosed. Neuroplacentology suggest that placental factors released into maternal and/or umbilical cord blood contribute to fetal brain development. Consistently, a preclinical inter-organ transcriptomic database revealed that PAE disrupts the expression ratio of angiogenic and inflammatory factors suggesting an angio-inflammatory response. This study aimed i) to assay, by multiplex immunoassay, angiogenic and inflammatory factors in maternal and umbilical cord blood from alcohol-consuming women and ii) to perform a maternofetal analysis according to neonatal sex. Afterwards, dysregulated factors from mothers who gave birth to females or males were submitted to *STRING* and *ShinyGO* analyses. Results showed that PAE differently altered the distribution profiles of dysregulated angiogenic and inflammatory factors in maternal and umbilical cord blood. Moreover, sex-specific differences were observed, with 36% of dysregulated proteins specific to males, 48% to females, and 16% common to both. *STRING* analysis revealed robust functional protein-protein interactions linking together inflammatory and angiogenic clusters while the *ShinyGO* analysis identified enriched pathways related to vascular shear stress. These findings provide the first maternofetal analysis of combined angiogenic and inflammatory factors from alcohol-consuming mothers.

## Introduction

Prenatal alcohol exposure (PAE) is a leading cause of neurodevelopmental disorders and secondary disabilities (1). A diagnosis of Fetal Alcohol Syndrome (FAS), the most severe expression of Fetal Alcohol Spectrum Disorders (FASD), can be established perinatally based on characteristic facial dysmorphologies, microcephaly, low birth weight and evidence of alcohol exposure (2). However, majority of neonates affected by FASD do not exhibit the distinctive craniofacial features associated with FAS and many infants are late- or mis-diagnosed (3, 4). Improving the early diagnosis of FASD could offer an additional opportunity for recovery. During the developmental window of maximum brain plasticity in the first three years of a child’s life, it has been shown that the earlier a neurodevelopmental disorder is identified, the earlier appropriate rehabilitation is implemented, the earlier parents are enrolled in parenting guidance programs, the earlier the child is integrated into a group setting, the better the recovery of an appropriate neurodevelopmental trajectory, and the better the prevention of secondary neurodisabilities and psychiatric disorders during preadolescence and adolescence (5–9).

Global epidemiological studies indicate that, despite active primary prevention, the proportion of women consuming alcohol during pregnancy as well as prevalence of FASD remain high (10). For example, the global prevalence of FASD among children and youth in the general population was estimated to be 7.7 per 1000 people with world region disparities (10). In Europe, FAS prevalence is estimated between 0.5–3.7 per 1000 people with country variations (11) while prevalence of FASD is estimated to be up to 10-fold higher (9, 12).

In cases of suspicion of alcohol consumption during pregnancy, several approaches including screening questionnaires and biomarker assays are used to confirm the fetal alcohol exposure (13, 14). Ideally, preconception consultation by healthcare professionals should be done as preventative screening to alert the future parents to the risks of alcohol exposure for the fetus.

However, to date no clear threshold of exposure at risk for the fetus has been established. While heavy alcohol exposure leads to teratogenic effects (15), several clinical and pre-clinical studies also reported that moderate doses and/or episodic consumption patterns can also disrupt the neurodevelopment of the fetus (16, 17). Conversely, not all exposed fetuses will necessarily develop clinical manifestations (18). This illustrates the complex and multifactorial nature of the alcohol toxicity for the fetus. Indeed, alcohol effects express differently according to numerous variables such as the period of pregnancy, the duration and dose of exposure, the maturation degree of a given cell type (19) or a given brain structure (20), poly-intoxication with other drugs such as tobacco (21) as well as individual genetic polymorphisms (22). Moreover, increasing evidence support sex-specific effects of PAE resulting in different impacts on gene expression patterns (23), brain region development (24) and integrative functions (25). This complex etiology of FASD extends before conception and beyond fetal life as alcohol exposure induces epigenetic modifications impacting germ cells of the future parents (26) and lasting postnatal effects on brain function and children’s behavior (27).

Recent studies related to the emerging field of neuroplacentology support that circulating agents released by the placenta can be detected in the maternal blood and/or umbilical cord blood and contribute to the neurodevelopment of the fetus (28–30). A recent inter-organ transcriptomic database of the “Placenta-Fetal cortex” signature of PAE revealed that alcohol disrupted the expression ratio of different genes involved in angiogenesis and inflammation (31, 23). Moreover, functional validation experiments consisting in a targeted repression of placental angiogenic factors i.e. placental-growth factor (PLGF) (32) and CD146, an adhesion molecule of the immunoglobulin superfamily mainly expressed in endothelium (30), revealed neurovascular abnormalities in the fetal brain. Taken together, these data suggest that circulating agents with angiogenic and inflammation properties may reflect PAE-induced dysfunctions contributing to neurovascular impairments. Such dysregulated factors could be representative of common signaling pathways able to refine the understanding of FASD pathophysiology. Based on previous pre-clinical transcriptomic and functional studies (23, 30, 31), the aims of the present study were i) to assay a panel of angiogenic and inflammatory factors in maternal blood and umbilical blood from alcohol-consuming women and ii) to perform a maternofetal analysis of dysregulated signaling pathways considering the sex of neonates.

## Results

The study included five clinical centers with 59 enrolled mothers and their neonates. Detailed information about the mothers (**Tables 1** and **2**) and their neonates (**Table 3**) was obtained from medical records at the time of delivery (**Supplementary Table 1**).

**Table 1.** Maternal characteristics across control and alcohol consuming groups. Continuous data are presented as: mean (Standard Error of Mean). Categorical variables are presented as: number (%). Comparisons between non-consuming and alcohol-consuming pregnant women were performed using Mann-Whitney *U* test for non-normal distributed continuous variables and Fisher’s exact test for categorical variables. Statistical significance was considered at p<0.05* and p<0.0001****. Complications were reported for 25 women including 13 pregnancy complications (n): cholestasis (1 control, 1 alcohol), gestational diabetes (5 control, 4 alcohol), intrauterine growth retardation (1 control, 1 alcohol). Drugs Intake (n): marijuana (15), heroin (1), cocain (3). SEM. Standard Error Mean; BMI. body mass index.

| Groups<br>n (%) | All<br>59 (100) | Control<br>31 (52.54) | IUAЕ<br>28 (47.46) | P<sub>Value</sub><br>Control vs<br>IUAЕ |
| --- | --- | --- | --- | --- |
| **Maternal/pregnancy variables** |  |  |  |  |
| Age (years) mean (SEM) | 30.07 (0.88) | 29.35 (1.01) | 30.86 (1.49) | 0.3236 |
| Pre-pregnancy BMI (kg/m<sup>2</sup>) mean (SEM) | 24.67 (0.91) | 26.53 (1.47) | 22.61 (0.89) | **0.0219\*** |
| Tertiary education n (%) |  |  |  |  |
| Yes | 20 (33.89) | 15 (48.38) | 5 (17.86) | **0.0396\*** |
| No | 29 (49.15) | 12 (38.71) | 17 (60.71) |  |
| Missing | 10 | 4 | 6 |  |
| Alcohol consumption habits n (%) |  |  |  |  |
| Chronic | 22 (37.29) | 0 (0) | 23 (82.14) |  |
| Acute | 6 (10.17) | 0 (0) | 6 (21.43) |  |
| Abstinence | 31 (52.54) | 31 (100) | 0 (0) |  |
| Tobacco use n (%) |  |  |  |  |
| Yes | 29 (49.15) | 5 (16.13) | 24 (85.71) | **<0.0001\*\*\*\*** |
| No | 30 (50.85) | 26 (83.87) | 4 (14.29) |  |
| Drugs used n (%) |  |  |  |  |
| Yes | 16 (27.12) | 0 (0) | 16 (57.14) | **<0.0001\*\*\*\*** |
| No | 43 (72.88) | 31 (100) | 12 (42.86) |  |
| Medication n (%) |  |  |  |  |
| Yes | 28 (47.46) | 11 (35.48) | 17 (60.71) | **0.0699** |
| No | 31 (52.54) | 20 (64.52) | 11 (39.29) |  |
| Complications n (%) |  |  |  |  |
| Yes | 25 (42.37) | 12 (38.71) | 13 (46.43) | **>0.9999** |
| No | 34 (57.62) | 19 (61.29) | 15 (53.57) |  |

**Table 2.** Alcohol consumption during pregnancy by period of exposure. . *Abbreviations*: T1-3. First to third trimester.

| IUAE group (n = 28) by periods of alcohol consumption |  |  |  |
| --- | --- | --- | --- |
| n | T1 | T2 | T3 |
| 17 | √ |  |  |
| 3 |  | √ |  |
| 8 | √ | √ | √ |

**Table 3.** Male and female measures of infants at birth. Continuous data are presented as: mean (Standard Error of Mean). Categorical variables are presented as number (%). Comparisons between non-exposed and *in utero* exposed to alcohol were performed using unpaired t-test for normally distributed continuous variables (male gestation length, male birth weight and length, female birth weight, and female head circumference). Mann-Whitney *U* test for non-normal distributed continuous variables and Fisher’s exact test for categorical variables. Statistical significance considered at p<0.05. IUAE: In Utero Alcohol Exposure; SEM: Standard Error Mean.

| Infant measures | All | Control | IUAE | P<sub>Value</sub> |
| --- | --- | --- | --- | --- |
| *Males n (%)* | 36 (100) | 19 (53) | 17 (47) |  |
| Gestation length (weeks) mean (SEM) | 37.39 (0.42) | 37.58 (0.47) | 37.18 (0.72) | 0.6379 |
| Birth weight (g) mean (SEM) | 2755 (114.5) | 2968 (152.6) | 2516 (157.3) | **0.0473\*** |
| Head circumference (cm) mean (SEM) | 33.03 (0.36) | 33.50 (0.50) | 32.56 (0.52) | 0.1531 |
| Birth length (cm) mean (SEM) | 47.41 (0.61) | 48.17 (0.74) | 46.56 (0.98) | 0.1952 |
| Preterm <37 weeks n (%) | 11 (30.6) | 5 (26.3) | 6 (12.8) | 0.7206 |
| Birth weight <10<sup>th</sup> centile n (%) | 10 (27.8) | 3 (15.8) | 7 (41.2) | 0.1394 |
| *Females n (%)* | 23 (100) | 12 (52) | 11 (48) | P<sub>Value</sub> |
| Gestation length (weeks) mean (SEM) | 39.00 (0.42) | 39.17 (0.58) | 38.83 (0.63) | 0.5662 |
| Birth weight (g) mean (SEM) | 3117 (123.3) | 3316 (193.2) | 2935 (144.3) | 0.1247 |
| Head circumference (cm) mean (SEM) | 33.87 (0.36) | 33.92 (0.42) | 33.82 (0.63) | 0.8958 |
| Birth length (cm) mean (SEM) | 48.35 (0.44) | 49.17 (0.52) | 47.45 (0.65) | **0.0398\*** |
| Preterm <37 weeks n (%) | 3 (8.3) | 1 (5.3) | 2 (11.8) | 0.5901 |
| Birth weight <10<sup>th</sup> centile n (%) | 5 (13.9) | 1 (5.3) | 4 (23.5) | 0.1550 |

### Maternal characteristics

Among the 59 participants, the mean maternal age was 30 +/− 0.88 years, and the mean pre-pregnancy BMI fell within the healthy range (**Table 1**). Tertiary education was reported by 20 participants (33.89%). Nearly half of the participants reported smoking during pregnancy (49.15%), 16 reported drug use (27.12%), and 28 reported taking medication (47.46%; **Table 1**). Complications were documented in 25 women (42.37%). Thirteen were directly associated to pregnancy and consisted in cholestasis (2), gestational diabetes (9) and intrauterine growth retardation (2; **Supplementary Table 1**). Comparison of maternal characteristics revealed no significant differences between the control and alcohol-consuming groups regarding maternal age (p = 0.3236) and pregnancy complications (p > 0.9999; **Table 1**). A trend toward medication use was noted (p = 0.0699; **Table 1**). Although pre-pregnancy BMI remained within the healthy range overall, it was significantly lower in the alcohol-consuming group compared with the control group (p = 0.0219). Tobacco use was also significantly more prevalent in the alcohol-consuming group (p < 0.0001). Drug use was absent in controls but present in over half of the alcohol-consuming women (57.1%; p < 0.0001), mainly marijuana (n = 15), with fewer reporting heroin (n = 1) or cocaine (n = 3; **Supplementary Table 1**). Finally, special attention was paid to alcohol consumption habits. Among the 28 alcohol-consuming pregnant women, 22 reported chronic alcohol consumption, while 6 reported acute consumption.

### Periods of alcohol consumption

The distribution of alcohol consumption during each stage of pregnancy is shown in **Table 2**. Among all women who consumed alcohol during pregnancy, 17 reported alcohol consumption during the first trimester, 3 during the second trimester and 8 reported alcohol consumption throughout all pregnancy (**Table 2**).

### Newborn characteristics

The results are presented in **Table 3**. The cohort included 36 male and 23 female neonates. Mean gestational age was 37.39 +/− 0.42 weeks for males and 39 +/− 0.42 weeks for females (**Table 3**). Preterm birth occurred less frequently among female neonates (8.3%) than among males (30.6%). Overall, mean birth weight fell between the 25th and 50th percentiles; however, birth weight below the 10th percentile was observed in 27.8% of male neonates and 13.9% of female neonates. A significant reduction in birth weight was identified in male neonates from the *in utero* alcohol exposed (IUAE) group compared with controls (p = 0.0473; **Table 3**). Among female neonates, a significant reduced birth length was noted in the IUAE group (p = 0.0398; **Table 3**). No significant differences were detected in head circumference (p = 0.1531 for males; p = 0.8958 for females) or in preterm birth rates between infants born to mothers who consumed alcohol during pregnancy and those born to mothers who did not (p = 0.7206 for males; p = 0.5901 for females; **Table 3**).

### Protein levels on sera from maternal blood

The expression levels of 42 proteins associated with angiogenesis and inflammation were quantified in the blood of 31 pregnant women from the control group and 28 mothers who consumed alcohol during pregnancy. The mean serum levels of each protein are shown in **Table 4a**. When tested for statistical significance without sex consideration, 10 of the analyzed proteins (bFGF, GM-CSF, IL-1β, IL-6, IL-8, IL-10, IL-12, IL-12p70, slCAM-1, TNFα) were significantly different between the two groups (**Table 4a**). Five of the analyzed proteins (bFGF, IL-8, IL-12p70, sICAM-1, TNFα) exhibited higher expression in the alcohol group and 5 (GM-CSF, IL-1β, IL-6, IL-10, IL-12) were lower (**Table 4a**). A trend was found for 6 proteins (CD146, TIE2, VEGFc, Eotaxin 3, IL-4, IL-13). Three factors (CD146, Eotaxin 3, IL-13) were increased in the alcohol group and 3 (TIE2, VEGFc, IL-4) were decreased (**Table 4a**). When considering the sex of neonates, in the blood of mothers who gave birth to a male, 8 of the 42 analyzed proteins (CD146, VEGF-R2, bFGF, GM-CSF, IL-6, IL-10, IL-13, sICAM-1) were significantly modified (**Table 4a**). Among them, 6 (CD146, VEGF-R2, bFGF, GM-CSF, IL-13, sICAM-1) showed higher expression in the alcohol group and 2 (IL-6, IL-10) were decreased (**Table 4a**). A trend was found for 3 proteins (VEGFc, IL-1β, IL-12) whose levels were decreased in the alcohol group (**Table 4a**). Considering mothers who gave birth to a female, 1 protein (TNFα) was significantly different between the two groups and its level was higher in the alcohol group (**Table 4a**). A trend was found for 7 proteins (TIE2, VEGF-R2, CRP, IL-8, IL-12, IL-12p70, IL-16). Levels of TIE2, VEGF-R2, CRP and IL-12 were decreased in the alcohol group whereas IL-8, IL-12p70 and IL-16 were increased (**Table 4a**).

**Table 4a.**
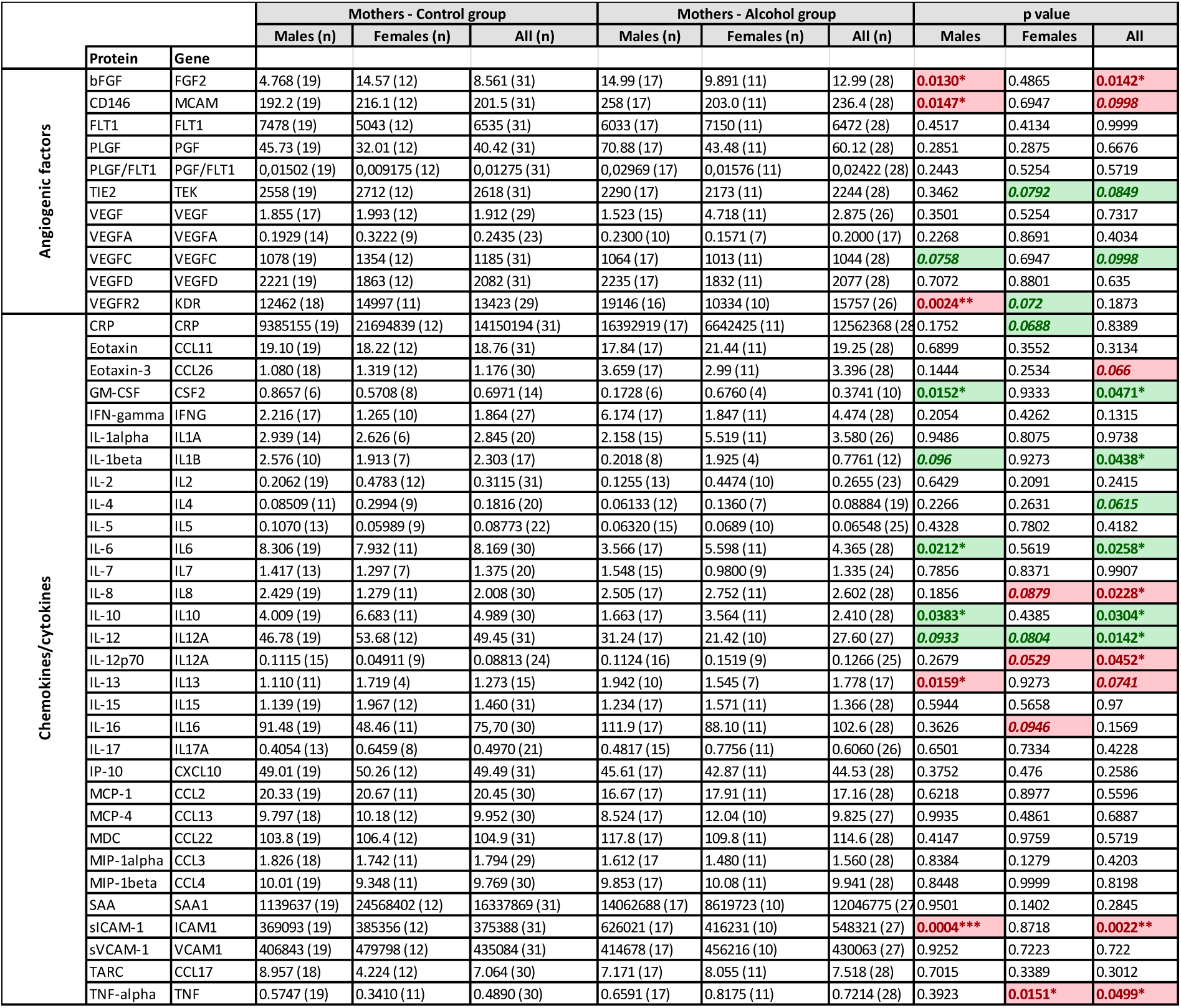
Mean serum levels of angiogenic and inflammatory factors in the blood of mothers from control and alcohol groups. In a given group (control and alcohol), data are presented as: mean (n) considering all mothers, mothers who gave birth to a male and mothers who gave birth to a female. For each factor, the detailed graph is presented in Supplementary Figure 1. Data are expressed as pg/ml excepted for the PLGF/VEGFR1 ratio. Comparisons between control and alcohol groups were performed using Mann-Whitney *U* test. Statistical significance is considered at p<0.05*, p<0.01**, p<0.001*** and a trend (italic values) is considered at 0.05<p< 0.1. Values highlighted in red and green indicate increased and reduced levels in the alcohol *versus* control groups, respectively.

### Protein levels on sera from umbilical cord blood

Protein levels from the angiogenesis and inflammation panels were also quantified in the umbilical cord blood of control and IUAE neonates. The mean serum levels of each protein are shown in **Table 4a**. When tested for statistical significance without sex consideration, levels of 5 proteins (TIE2, IFNɣ, IL-1β, IL-6, IL-13) were significantly modified between the two groups (**Table 4a**). Three of the analyzed proteins (IFNɣ, IL-6, IL-13) exhibited higher levels in the alcohol group and 2 (TIE2, IL-1β) were lower (**Table 4a**). A trend was found for 5 proteins (VEGF-D, IL-10, IL-12p70, IL-15, IL-17). One factor (IL-15) was increased in the alcohol group and 4 (VEGF-D, IL-10, IL-12p70, IL-17) were decreased (**Table 4a**). Considering male neonates, 3 of the analyzed proteins (VEGF-R2, IL-1β, IL-13) were significantly modified in the IUAE group (**Table 4a**). VEGF-R2 and IL-13 exhibited higher levels in the alcohol group, whereas IL-1β was lower. A trend was found for 3 proteins (CRP, IFNɣ, slCAM-1). All of them presented higher levels in the alcohol group (**Table 4a**). Considering female neonates, 6 of the 42 analyzed proteins (bFGF, IL-7, IL-12p70, IL-17, MCP4, MIP1β) were significantly modified in the IUAE group (**Table 4a**). All of them were decreased in the alcohol group (**Table 4a**). A trend was found for 4 proteins (TIE2, VEGFA, Eotaxin, TARC). All of them were decreased in the alcohol group (**Table 4a**).

**Table 4b.**
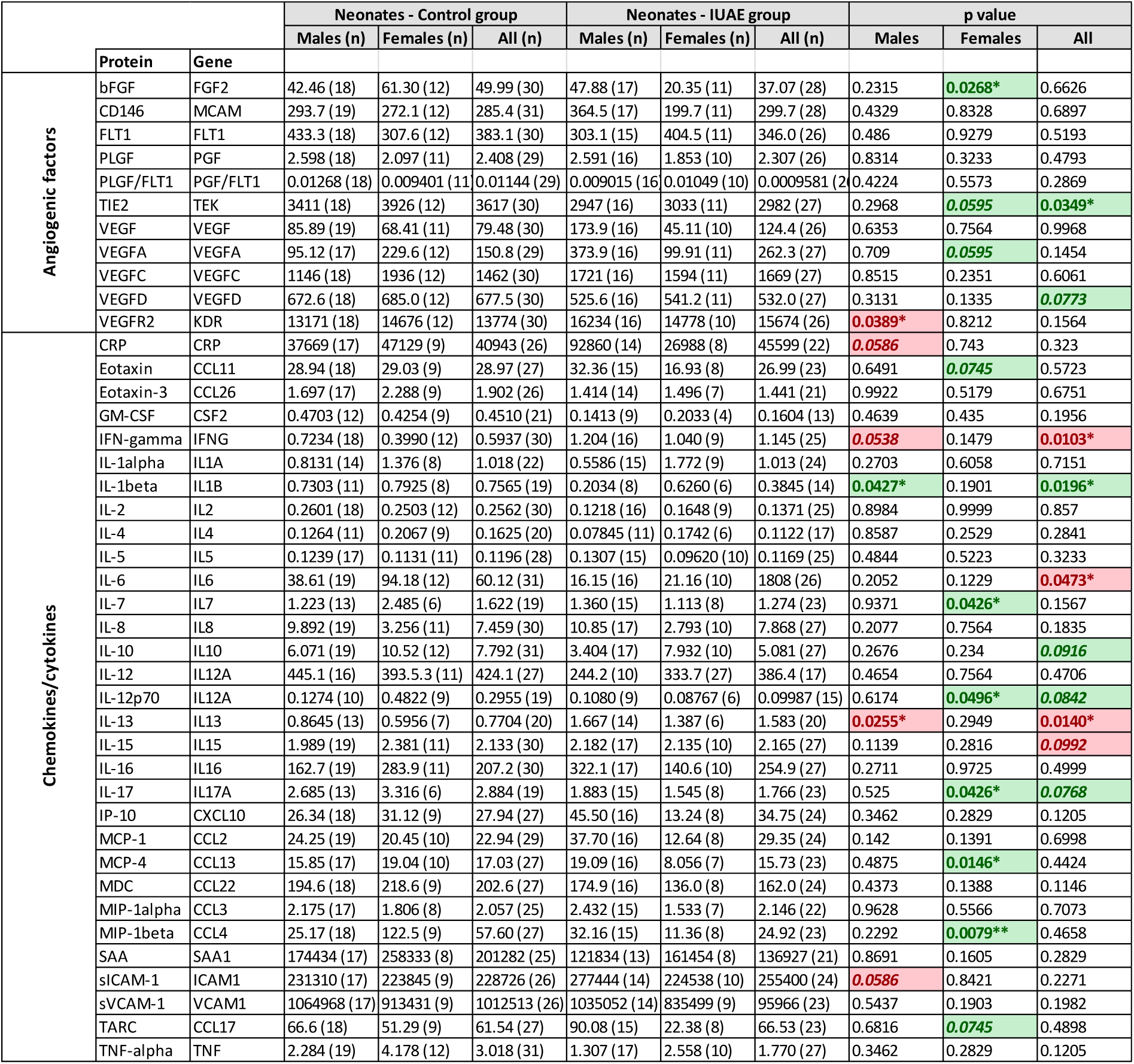
Mean serum levels of angiogenic and inflammatory factors in the umbilical cord blood of neonates from control and IUAE groups. Data are presented as: mean (n). In a given group (control and alcohol), comparisons were performed using Mann-Whitney *U* test in all neonates, males and females. Data are expressed as pg/ml excepted for the PLGF/FLT1 ratio. For each factor, the detailed graph is presented in Supplementary Figure 2. Statistical significance is considered at p<0.05*, p<0.01** and a trend (italic values) at 0.05<p< 0.1. Values highlighted in red and green indicate increased and reduced levels in the alcohol *versus* control groups, respectively. IUAE: In Utero Alcohol Exposure.

### Compilation of factors dysregulated by PAE in maternal blood and umbilical cord blood

Angiogenic and inflammatory factors modified between the control and the alcohol groups have been listed in **Table 5**. The comparison of factors impacted by alcohol consumption in the lists “*Mothers who gave birth to a male*” and “*Mothers who gave birth to a female*” showed that most dysregulated proteins were found in only one of the two lists: 36% were present in the “*Mothers who gave birth to a male*” list and 48% in the list “*Mothers who gave birth to a female*” (**Figure 1**). Only 16% (bFGF, CRP, IL12, VEGFR2) were common (**Figure 1**). Interestingly, among these common factors, bFGF, CRP and VEGFR2 were oppositely regulated when considering sex (**Table 5**). Altogether, these data support sex-dependent effects of alcohol consumption/exposure on circulating angiogenic and inflammatory factors.

**Figure 1.**
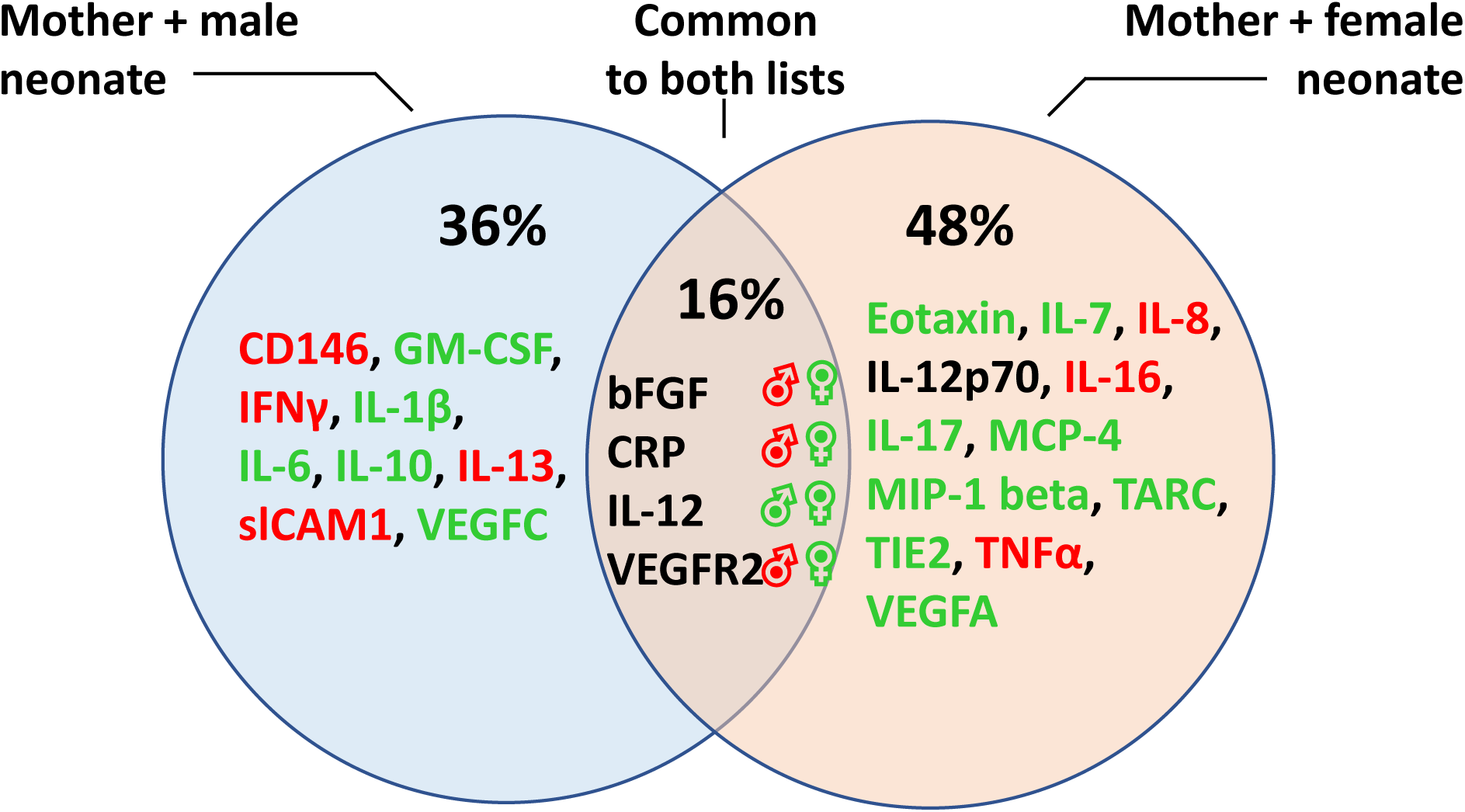
Venn diagram illustrating the distribution of dysregulated angiogenic and inflammatory factors considering the sex of neonates. Most angiogenic and inflammatory factors are differently distributed between mothers who gave birth to a male and mothers who gave birth to a female. Red and green colors indicate whether the factor was more or less expressed in the alcohol group. Regarding common factors, red and green colors indicate whether the factor was more or less expressed in the alcohol group depending of sex. Note that although common, several factors exhibit inverse regulation between the two groups.

**Table 5.** Lists of dysregulated angiogenic and inflammatory factors in control and alcohol groups. The lists compile factors with significant dysregulation (bold; p<0.05) and trend (italic; 0.05<p<0.1). Values highlighted in red and green indicate increased and reduced levels in the alcohol *versus* control groups, respectively. Column 1: factors from mothers who gave birth to a male; Column 2: factors from mothers who gave birth to a female; Column 3: factors from the umbilical cord blood of male neonates; Column 4: factors from the umbilical cord blood of female neonates; Column 5: pooled factors from columns 1 and 3; Column 6: pooled factors from columns 2 and 4.

| Mothers |  | Neonates |  | Mothers + Neonates |  |
| --- | --- | --- | --- | --- | --- |
| ♂ | ♀ | ♂ | ♀ | ♂ | ♀ |
| **bFGF / FGF2** |  |  | **bFGF / FGF2** | **bFGF / FGF2** | **bFGF / FGF2** |
| **CD146 / MCAM** |  |  |  | **CD146 / MCAM** |  |
|  | *CRP* | *CRP* |  | *CRP* | *CRP* |
|  |  |  | *Eotaxin/CCL11* |  | *Eotaxin/CCL11* |
| **GM-CSF / CSF2** |  |  |  | **GM-CSF / CSF2** |  |
|  |  | *IFN-gamma / IFNG* |  | *IFN-gamma / IFNG* |  |
| *IL-1beta / IL1B* |  | **IL-1beta / IL1B** |  | *IL-1beta / IL1B* |  |
| **IL-6** |  |  |  | **IL-6** |  |
|  |  |  | **IL-7** |  | **IL-7** |
|  | *IL-8 / CXCL8* |  |  |  | *IL-8 / CXCL8* |
| **IL-10** |  |  |  | **IL-10** |  |
| *IL-12 / IL12B* | *IL-12 / IL12B* |  |  | *IL-12 / IL12B* | *IL-12 / IL12B* |
|  | *IL-12p70* |  | **IL-12p70** |  | *IL-12p70* |
| **IL-13** |  | **IL-13** |  | **IL-13** |  |
|  | *IL-16* |  |  |  | *IL-16* |
|  |  |  | **IL-17 / IL17A** |  | **IL-17 / IL17A** |
|  |  |  | **MCP-4 / CCL13** |  | **MCP-4 / CCL13** |
|  |  |  | **MIP-1beta / CCL4** |  | **MIP-1beta / CCL4** |
| **sICAM-1 / CD54** |  | **sICAM-1 / CD54** |  | **sICAM-1 / CD54** |  |
|  |  |  | *TARC / CCL17* |  | *TARC / CCL17* |
|  | *Tie2 / TEK* |  | *Tie2 / TEK* |  | *Tie2 / TEK* |
|  | **TNFalpha / TNF** |  |  |  | **TNFalpha / TNF** |
|  |  |  | **VEGFA** |  | **VEGFA** |
| *VEGFC* |  |  |  | *VEGFC* |  |
| **VEGFR2 / KDR** | **VEGFR2 / KDR** | **VEGFR2 / KDR** |  | **VEGFR2 / KDR** | **VEGFR2 / KDR** |

### Protein-Protein Interaction (PPI) analysis of alcohol-dysregulated factors

A *STRING* analysis was performed to determine if the angiogenic and inflammatory factors found dysregulated in the lists “*Mothers who gave birth to a male*” and “*Mothers who gave birth to a female*” were or not functionally interacting (**Figure 2**). As predictable, the *STRING* analysis showed strong PPI enrichments within each cluster: inflammatory (blue area; **Figure 2A,C**) and angiogenic (pink area; **Figure 2B,C**). As an example, when considering the list “*Mothers who gave birth to a male*”, 6 high to highest confidence edges functionally linked IL6 with other inflammatory factors (**Figure 2A**). In the same way, in the group “*Mothers who gave birth to a female*”, 7 high to highest interactions linked TNFα with other inflammatory factors (**Figure 2B**). Interestingly, few but robust PPI confidence edges linked together some members of the inflammatory and angiogenic clusters. Indeed, when considering the group of “*Mothers who gave birth to a male”*, the *STRING* analysis revealed high confidence PPIs linking FGF2 and ICAM1 (angiogenic cluster) to the inflammatory factors IL-1β and IL6 (**Figure 2A**). In comparison, other members of the angiogenic cluster (MCAM, KDR, VEGFC) were poorly linked with the inflammatory cluster whereas they were strongly linked together (**Figure 2A**). Similarly, in the group of “*Mothers who gave birth to a female”*, FGF2 and VEGFA (angiogenic cluster) had robust PPI enrichments with TNFα and IL17 (inflammatory cluster; **Figure 2B**). Altogether, the *STRING* analysis identified robust sex-specific interactions linking together some inflammatory and angiogenic factors dysregulated by alcohol consumption during pregnancy suggesting their contribution in common signaling pathways.

**Figure 2.**
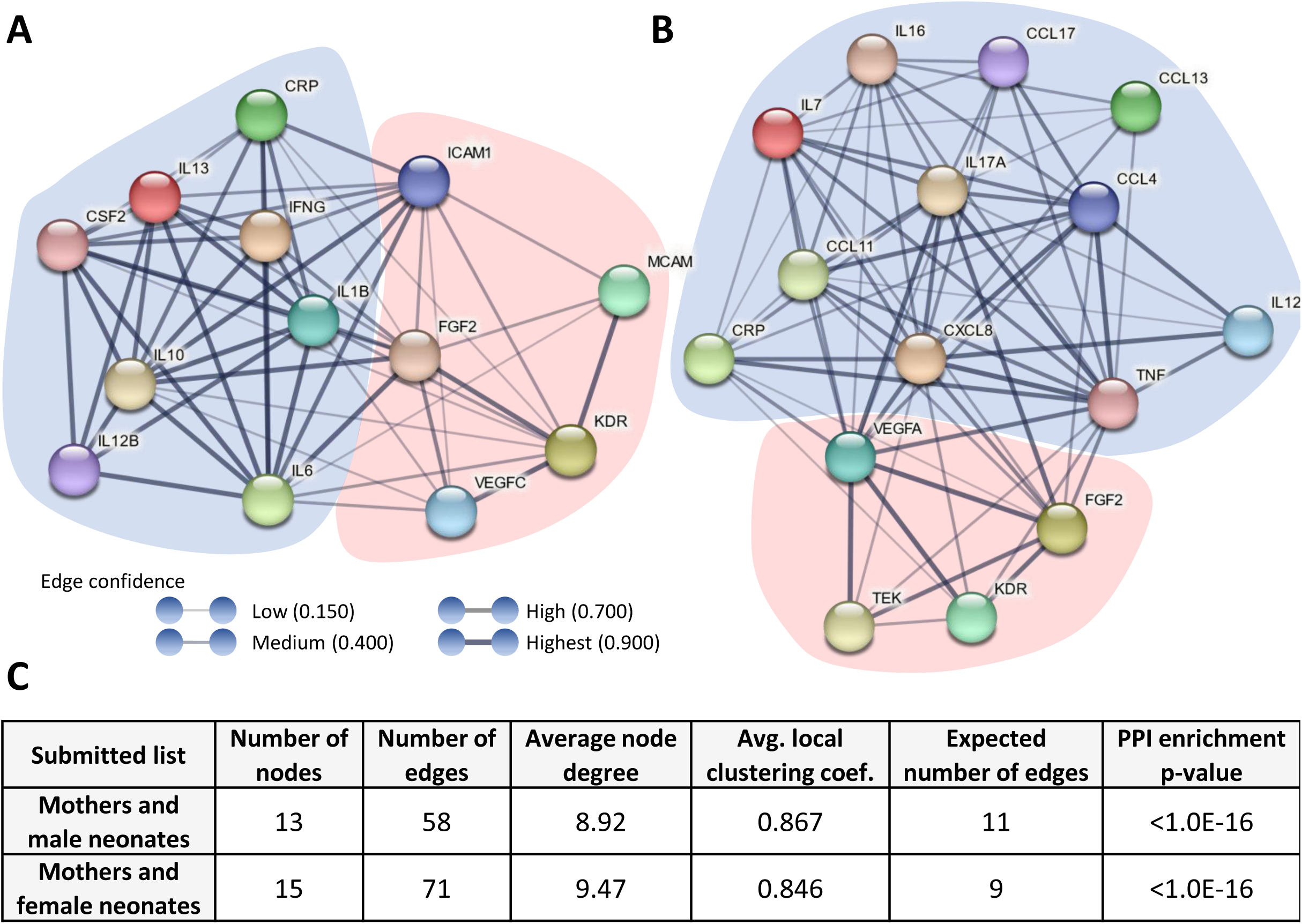
*STRING* analysis of the alcohol-dysregulated proteins between Control and Alcohol groups. **(A)** Visualization of protein-protein interactions between dysregulated inflammatory (blue cluster) and angiogenic (pink cluster) factors in the group “Mothers who gave birth to a male”. **(B)** Visualization of protein-protein interactions between dysregulated inflammatory (blue cluster) and angiogenic (pink cluster) factors in the group “Mothers who gave birth to a female”. Solid lines illustrate the protein-protein interactions (PPI). The width of edges represents the degree of confidence (the thickest lines, the highest confidence). **(C)** Table summarizing the network statistics for the two lists of proteins submitted to the *STRING* analysis. *Node*: protein; *Edge*: link between two proteins; *Average node degree*: number of interactions that a protein has on the average in the network. *Avg. local clustering coef.*: Index of clustering quality. A value below 1 suggests a good clustering. *Expected number of edges*: gives how many edges to expect if the nodes were to be selected at random. *PPI enrichment p-value*: A small PPI enrichment p-value indicate that nodes are not random and that the observed number of edges is significant.

### Signaling pathway analysis of alcohol dysregulated factors

In order to clarify if PPI enrichments found by the *STRING* analysis could be representative of dysregulated signaling pathways, the two lists “*Mothers who gave birth to a male*” and “*Mothers who gave birth to a female*” were submitted to a *ShinyGO* analysis (**Table 6**). As predictable, the *ShinyGO* analysis found significantly enriched signaling pathways related to inflammation such as the “*IL17 signaling pathway*” (KEGG #hsa04657; FDR=1.40E-08 in *Mothers who gave birth to a male*; **Table 6**) as well as to angiogenesis such as the “*VEGF signaling pathway*” (KEGG #hsa04370; FDR=2.1E-03 in *Mothers who gave birth to a female*; **Table 6**). Interestingly, the *ShinyGO* analysis also identified two enriched PAE-dysregulated signaling pathways linking together inflammatory and angiogenic factors *i.e*. the “*Fluid shear stress*” pathway (KEGG #hsa05418) in the group *Mothers who gave birth to a male* (FDR=4.60E-06) and in the group *Mothers who gave birth to a female* (FDR=4.4E-04; **Table 6**; **Figure 3A**) and the “*TNF signaling pathway*” (KEGG #hsa04668) in the group *Mothers who gave birth to a male* (FDR=3.8E-08; **Table 6**; **Figure 3B**). When considering the sex of neonates, the “*TNF signaling pathway*” was significantly enriched only in the group “*Mothers who gave birth to a male*” while the “*Fluid shear stress pathway*” was found significantly enriched in both groups (**Table 6**) suggesting a sex-different impact of PAE on angio-inflammatory processes.

**Figure 3.**
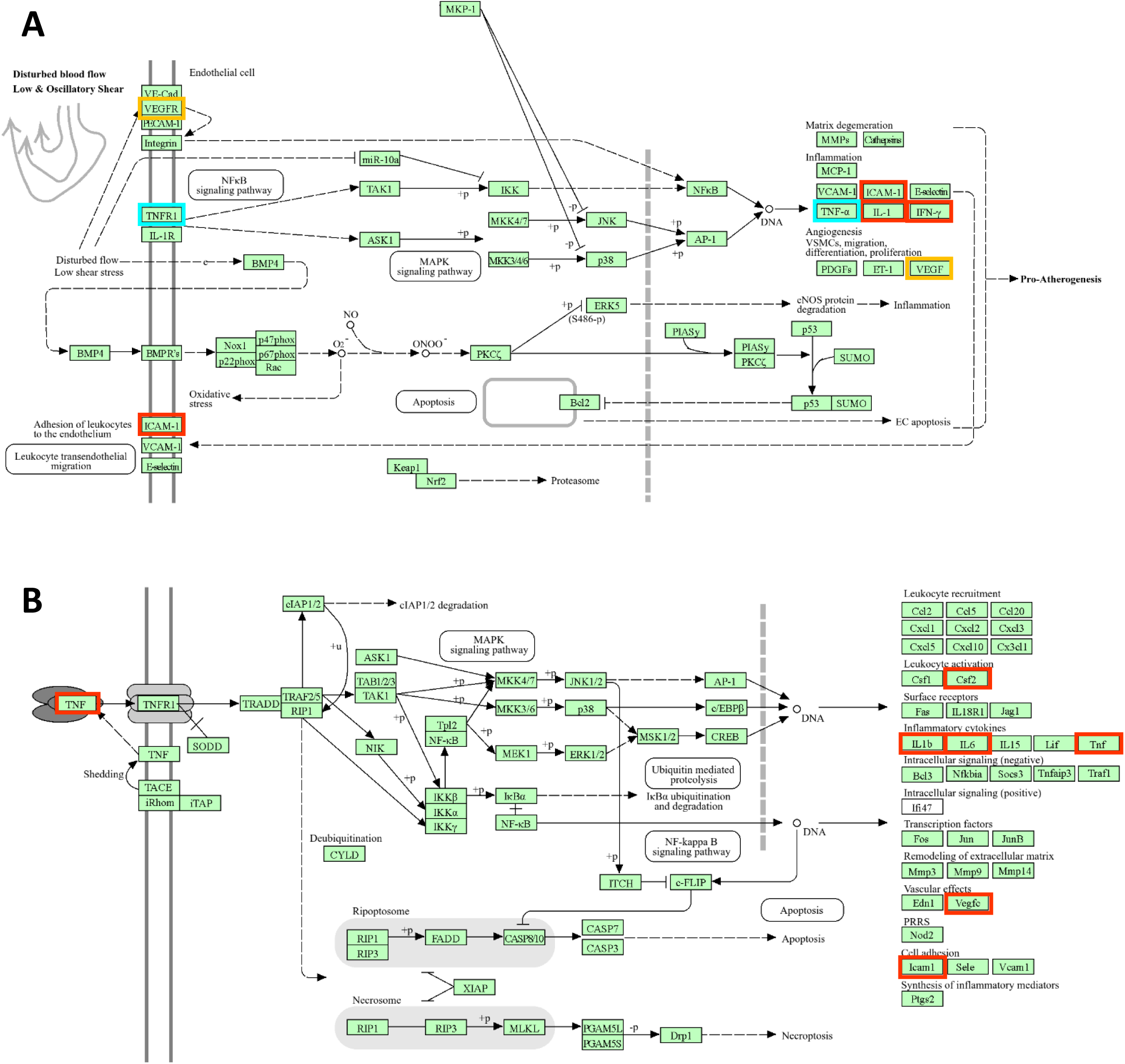
Visualization of cell signaling pathways significantly enriched and linking together dysregulated angiogenic and inflammatory factors. **(A)** The fluid shear stress signaling pathway. This pathway (#KEGG hsa05418) was found significantly enriched after *ShinyGO* analysis in the groups “*Mothers who gave birth to a male*” (FDR: 4.60E-06) and “*Mothers who gave birth to a female*” (FDR: 4.4E-04). Blue and red rectangles visualize in the pathway the factors found dysregulated in the groups “*Mothers who gave birth to a female*” (TNF-α, TNFR1) and “*Mothers who gave birth to a male*” (IL1, IFN-γ, ICAM-1), respectively. Yellow squares identify dysregulated factors in both groups (VEGF, VEGFR). (**B**) The TNF signaling pathway. This pathway (#KEGG hsa04668) was found significantly enriched after *ShinyGO* analysis in the group “*Mothers who gave birth to a male*” (FDR: 3.8E-08). Red rectangles visualize dysregulated factors (Csf2, IL1b, IL6, TNF, ICAM-1, VEGFc).

**Table 6.** Identification of enriched pathways related to inflammation and/or angiogenesis after *ShinyGO* analysis. Proteins found dysregulated between control and alcohol groups in the “*Mothers and male neonates*” and “*Mothers and female neonates*” list were submitted to a *ShinyGO* analysis. Five signaling pathways were found enriched. *KEGG pathway*: Name of the pathway; *KEGG number*: Reference of the *KEGG* pathway; *Enrichment FDR*: Reflects the statistical significance of the enrichment. An FDR adjusted p-value of 0.05 indicates that 5% of tested factors will result in false positives. *Number of genes*: number of genes coding the submitted proteins and present in the pathway. *Pathway genes*: total number of genes in a pathway. *Fold enrichment*: ratio of the percentage of genes in the list belonging to a pathway divided by the corresponding percentage in the background.

| Submitted list | KEGG pathways | KEGG number | Enrichment FDR | Number of genes | Pathway genes | Fold enrichment |
| --- | --- | --- | --- | --- | --- | --- |
| Mothers and male neonates | Cytokine-cytokine R interaction | hsa 04060 | 1.6E-09 | 7 | 297 | 42.2 |
|  | IL-17 signaling pathway | hsa 04657 | 1.40E-08 | 5 | 93 | 96.2 |
|  | TNF signaling pathway | hsa 04668 | 3.8E-08 | 5 | 119 | 75.2 |
|  | Fluid shear stress | hsa 05418 | 4.60E-06 | 4 | 141 | 50.8 |
| Mothers and female neonates | Cytokine-cytokine R interaction | hsa 04060 | 1.4E-12 | 9 | 297 | 50.3 |
|  | IL-17 signaling pathway | hsa 04657 | 4.4E-06 | 4 | 93 | 71.5 |
|  | Fluid shear stress | hsa 05418 | 4.4E-04 | 3 | 141 | 35.3 |
|  | VEGF signaling pathway | hsa 04370 | 2.1E-03 | 2 | 59 | 56.3 |

## Discussion

Building on preclinical models of FASD, the present manuscript provides the first clinical study co-analyzing inflammatory and angiogenic factors measured in maternal and umbilical cord blood from alcohol-consuming women during pregnancy. Based on the hypothesis that dysregulation of inflammatory and angiogenic factors in maternal blood and umbilical cord blood may reflect different sides of common signaling processes, the biostatistical analysis was performed by compiling together the factors found dysregulated by PAE in maternal blood and umbilical cord blood. Highlights of the study consist in the identification of sex-specific significant protein-protein interactions linking together maternofetal factors within an angio-inflammatory response.

### From pre-clinical models of FASD to FASD

Several pre-clinical studies evidenced that exposure of pregnant mice to alcohol is associated to altered expression of angiogenic factors in different organs such as the placenta (32, 35) or the fetal brain (19). Moreover, gain and loss of function experiments revealed that modified expression of angiogenic factors originating from the placenta such as IgF1 (28) and PLGF (32) alters the development of brain structures like the striatum (28) and the somatosensory cortex (32). In particular, it has been shown that placental repression of CD146, a pro-angiogenic factor, led to impaired differentiation of oligodendrocytes migrating along microvessels (30). Similarly, it has also been established that PAE modifies the expression of inflammatory factors such as IL-6 in the maternal blood (36) or IL-1β and TNF-α in the fetal brain (37, 38). These data obtained in FASD models are consistent with several publications performed in human which showed that alcohol consumption during pregnancy modified the expression of angiogenic factors such as PLGF with sex-dependent effects in the placenta (39) as well as VEGF, MMP9 or angiogenin in the umbilical cord blood (40). Alcohol consumption during pregnancy has also been shown to modify the expression of inflammatory factors such as IL-1β, IL-6, IL17d or TNFα in the mother, the placenta or the fetus (41, 42). Although several recent studies highlighted the contribution of peripheral angiogenic factors on fetal brain development (28, 30) no joint analysis of the effects of PAE on angiogenic and inflammatory protein levels has ever been conducted. The present study revealed that several angiogenic (bFGF, CD146, VEGFA, VEGFC, VEGFR2…) and inflammatory (CRP, IL1-β, IL-4, IL6, TNF-α…) factors were dysregulated by alcohol consumption in maternal blood and/or umbilical cord blood. Different hypotheses could be raised to interpret these data: *i)* these dysregulated angiogenic and inflammatory factors in maternal and umbilical cord bloods are independently impacted by PAE and don’t share any common mechanistic interactions, or *ii)* supported by recent advances regarding neuroplacentology, these factors dysregulated in the maternal and umbilical cord bloods would reflect different aspects of common “angio-inflammatory” processes.

### Linking inflammation and angiogenesis in a context of alcohol consumption during pregnancy

In order to check the two hypotheses mentioned above, a biostatistical analysis was carried out and consisted in grouping together dysregulated factors from maternal blood and umbilical cord blood and performing a joint analysis of the angiogenic and inflammatory clusters. When considering a given cluster separately (angiogenic or inflammatory), the *STRING* analysis revealed substantial protein-protein interactions supporting that, as already shown in the literature, PAE exerts inflammatory (42, 43) and angiogenic (40, 44, 45) effects. Consistently, the *ShinyGO* analysis identified enriched *KEGG* pathways specifically related to inflammation (IL-17) and angiogenesis (VEGF). When considering inter-cluster enrichments, the *STRING* analysis identified few but robust PPI linking together different angiogenic factors (FGF2, ICAM1, VEGFA) with inflammatory factors (IL-6, IL-1β, TNFα). Analysis of the literature suggested such interactions in other pathophysiological contexts. In adult rats, Barney and co-workers showed that peripheral injection of ethanol increased IL-6 and FGF2 expression in the hippocampus (46) while in an animal model of alcohol-induced gastric ulcer, the dysregulation of cytokines (TNFα, IL-1 β and IL-6) was associated to altered expression of the adhesion protein ICAM1 (47). In human, Bodnar and co-workers showed that alcohol consumption altered blood levels of immune factors (IFN-ɣ, IL-10, TNF-β, TNF-α, and CRP) as well as members of the VEGF family (48). In the present study, the *ShinyGO* analysis refined the *STRING* PPI analysis by identifying significantly enriched signaling pathways linking together inflammatory and angiogenic factors: the *TNFα pathway* (#hsa 04668) and the *Fluid shear stress pathway* (#hsa 05418). It is interesting to note that, in adult cardiovascular diseases, disturbed blood flow contributes to a vasculo-inflammatory response in which TNFα plays a central role, leading to dysregulated expression of endothelial adhesion molecules such as ICAM-1 and VCAM-1 (49). Considering the present study and recent data from Steane and co-workers who showed that, in alcohol-consuming pregnant women, the feto-placental blood flow is disturbed (39), it is tempting to speculate that a vasculo-inflammatory response could contribute in the vascular impairments described in human placentas (32) and FAS fetuses (50).

### Sex-specificities

Several pre-clinical and clinical studies support that the effects of PAE are differently expressed between females and males. In a mouse model of FASD, an inter-organ transcriptomic analysis between the placenta and the fetal cortex showed that PAE differentially impacted the expression of neurotrophic factors between females and males (23). Similarly, it was shown in rats, that PAE durably and sex-dependently impacted the immune profile of the offspring (51). In human, few studies investigated the inflammatory and angiogenic effects of PAE by considering separately males and females. However, Steane and co-workers showed that in male placentas, PAE reduced the expression of PLGF mRNA, whereas no effect was found in female placentas (39). At a morphometric level, MRI analyses of brain structures showed sex-specific differences between FASD and control subjects (52, 53). Moreover, Long and co-workers showed sex-specific associations between gray matter volumes and executive functions (24). Finally, sex-related differences have been described regarding the clinical expression of FASD across lifespan (25). In the present study, the alcohol-induced dysregulations of angiogenic and inflammatory factors were analyzed considering separately mothers who gave birth to a female or to a male. In both sexes, PAE had a significant impact on immune and angiogenic factors. However, numerous dysregulated factors appeared different and, when common, oppositely regulated between males and females. For example, while bFGF was increased by PAE in the group “*Mothers who gave birth to a male*”, it was decreased in the group “*Mothers who gave birth to a female*”. Similarly, whereas CRP and VEGFR2 levels were impacted by PAE, they were oppositely regulated when the neonate was a female or a male. Altogether, these data suggest that, as supported by pre-clinical studies (36), alcohol appears to induce a sex-specific angio-inflammatory response in human during pregnancy.

### Limitations and perspectives. Design of angiogenic and immune factors

Forty-two angiogenic and immune factors were quantified in the maternal blood and umbilical cord blood from control and alcohol-consuming mothers by using V-Plex panels validated for clinical research. While providing a large screening of inflammatory and angiogenic factors, the use of such panels remains non-exhaustive. This point raises a risk of missing angiogenic/inflammatory interactions. Such limitation could be reduced by using omics approaches as recently done in a preclinical model of FASD (23).

### Small group size

Strength and originality of the present study consist in *i)* considering that, even if different, dysregulated factors in maternal blood and umbilical cord blood could represent the expression of common mechanistic processes, and *ii)* discriminating the mothers who gave birth to a female and to a male. Such strategy allowed to identify significantly enriched protein-protein interactions and signaling pathways. However, the small sizes of the groups constitute a weakness of the study (31 inclusions in control group and 28 inclusion in alcohol-consuming women). Indeed, a higher number of inclusions would be able to provide a more accurate vision of the angio-inflammatory hypothesis. In perspective, the PAE-induced angio-inflammatory hypothesis could be functionally validated by returning to FASD preclinical models as recently done for the functional validation of neuroplacental interactions (30).

### Prenatal exposure to other toxics

As described in several clinical protocols (39, 54), maternal characteristics from the present study showed that alcohol-consuming women also consume other toxic substances; tobacco being the most common. Moreover, several publications reported that maternal smoking promotes inflammation (55) and disrupts angiogenic balance (56). Consequently, it is important to keep in mind that polyintoxication behaviors constitute a unifying mechanism for adverse neurodevelopmental outcomes associated with fetal programming (57). However, translational studies such as those from Lecuyer and co-workers which compared placenta/brain phenotypes in human FASD with placenta/brain phenotypes from a mono-intoxication mouse model of FASD constitutes a promising approach to decipher the contribution of PAE on vasculo-inflammatory processes (32).

In conclusion, this clinical study provides the first joint analysis of angiogenic and inflammatory factors in maternal and umbilical cord blood from women who consumed alcohol during pregnancy. Biostatistical analysis revealed sex-specific PAE signatures of dysregulated angiogenic and inflammatory factors. Protein-protein interaction analysis revealed that several of these angiogenic and inflammatory factors exhibit strong functional interactions and belong to pathophysiological angio-inflammatory responses already characterized in adult diseases and related to vascular shear stress. These original findings can be associated with recent preclinical advances in neuroplacentology which have revealed a contribution of circulating placental angiogenic factors to the neurovascular development of the fetal brain.

## Methods

### Study design

The multicentric AlcoBrain protocol was conducted from 2017 to 2021 under the supervision of Pr S. Marret (Charles Nicolle Hospital, Rouen, France) and involved four additional clinical centers (Dr A.L. Duigou, Morvan Hospital, Brest, France; Dr V. Datin-Dorriere, Caen University Hospital, Caen, France; Dr H. Bruel, Le Havre Hospital, Le Havre, France; Pr V. Biran, Robert Debre Hospital, Paris, France). The AlcoBrain protocol was registered at the National Agency for the Safety of Medicines and Health Products (ANSM) under the identification number RCB: 2016-A01563-48. On January 27, 2017, the AlcoBrain protocol received approval from the Ethical committee “*Comité de Protection des Personnes Nord-Ouest I*” of the Charles Nicolle Hospital, Rouen, France (n°CPP: 02-021-2016) for the prospective recruitment of 66 pregnant women (**Figure 4**). Recruitment was performed during antenatal consultations or appointments in the five clinical centers. Written consent was obtained by an investigator on the day of delivery, prior to any research-related procedures. For the child’s participation, consent from both the father and the mother was obtained. Detailed information about the mothers and their infants was obtained from medical records at the time of delivery (**Supplementary Table 1**).

**Figure 4.**
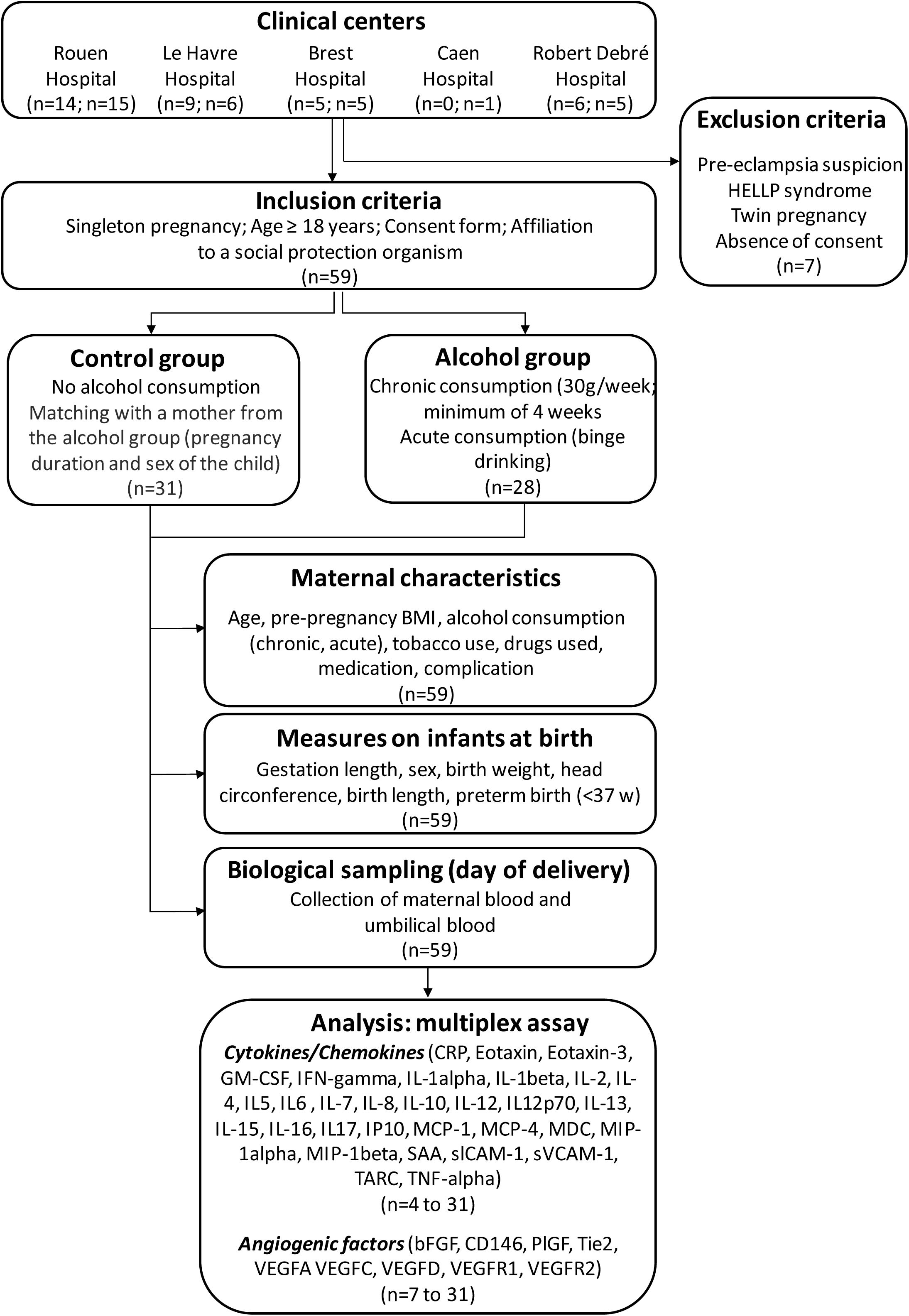
Flow chart of the study. Indicators mention the number of centers involved in the recruitment, the number of participants enrolled (exclusions and final number included), the maternal and infant characteristics, and the biochemical analysis perfomed on maternal and umbilical bloods. Abbreviations: bFGF. basic Fibroblast Growth Factor; CD146. Cluster of Differenciation 146; CRP. C-Reactive Protein; GM-CSF. Granulocyte Macrophage Colony-Stimulating Factor; IFN-γ. Interferon-γ; IL-1α/β. Interleukine 1α/β; IL-2/4/5/6/7/8/10/12/13/15/16/17. Interleukine 2/4/5/6/7/8/10/12/13/15/16/17; IP-10. Interferon γ-inducible protein 10; MCP-1/4. Monocyte Chemoattractant Protein-1/4; MDC. Macrophage-Derived Chemokine; MIP-1α/β. Macrophage Inflammatory Protein-1α/β; PlGF. Placental Growth Factor; SAA. Serum Amyloid A; sICAM-1. soluble Intercellular Cell Adhesion Molecule-1; sVCAM-1. soluble Vascular Cell Adhesion Molecule-1; TARC. Thymus and Activation Regulated Chemokine; Tie2. Tyrosine kinase with immunoglobulin and epidermal growth factor homology domains 2; TNF-α. Tumor Necrosis Factor-α; VEGFA/C/D. Vascular Endothelial Growth Factor A/C/D. VEGFR1/2. Vascular Endothelial Growth Factor Receptor 1/2.

### Participants

To be included in the study, the participants had to be carrying a singleton pregnancy, be at least 18 years old and be affiliated to a social protection organism (**Figure 4**). Among the 66 pregnant women, 7 who developed pre-eclampsia, HELLP syndrome, or had a twin pregnancy were excluded (**Figure 4**). In addition, women who were unable to provide informed consent were excluded. For the control group, 31 women were included (**Figure 4**). They must not have consumed alcohol throughout the pregnancy and were matched with a woman from the alcohol group. Women in the control group were progressively included as women who had consumed alcohol during pregnancy were enrolled. For the alcohol group, 28 women were included (**Figure 4**). Participants should have had documented alcohol consumption during pregnancy, including either chronic consumption of at least 30 grams of alcohol per week or acute consumption (binge drinking; **Figure 4**). All included mothers received care at the maternity units of the study centers. Common clinical information about the mothers such as age, pre-pregnancy body mass index (BMI), tobacco use, drug use, medication use, and pregnancy complications, were collected (**Supplementary Table 1**). A pediatric examination at birth, including somatometric measurements such as height, weight, and head circumference, were performed (**Figure 4**).

### Biological sampling

Two tubes of maternal blood (a 2 mL dry tube and a 3 mL EDTA tube) were collected on the day of delivery. Two tubes of blood from the umbilical cord (a 2 mL dry tube and a 3 mL EDTA tube) were also collected (**Figure 4**). All samples were anonymized, centrifuged, the supernatants were aliquoted, and then stored at −20°C until analysis. Serum levels of angiogenic factors and chemokines/cytokines were measured in maternal and umbilical cord blood from both groups.

### Electrochemiluminescence Immunoassays (ECLIAs)

A multiplex assay using the V-Plex Vascular Injury Panel 2 Human Kit (Cat # K15198D, MSD, Rockville, Maryland, USA), the V-Plex Angiogenesis Panel 1 Human Kit (Cat # K15190D, MSD), the V-Plex Chemokine Panel 1 Human Kit (Cat # K15047D, MSD), the V-Plex Cytokine Panel 1 Human Kit (Cat # K15050D, MSD) and the V-Plex Proinflammatory Panel 1 Human kit (Cat # K15049D, MSD) were carried out to analyze protein levels in pg/mL of bFGF, CD146, CRP, Eotaxin, Eotaxin-3, GM-CSF, IFN-γ, IL-1α, IL-1β, IL-2, IL-4, IL-5, IL-6, IL-7, IL-8, IL-10, IL-12, IL-12p70, IL-13, IL-15, IL-16, IL-17, IP10, MCP-1, MCP-4, MDC, MIP-1α, MIP-1β, PLGF, SAA, sICAM-1, sVCAM-1, TARC, TIE2, TNFα, TNFβ, VEGFA, VEGFC, VEGFD, VEGFR1 and VEGFR2 according to manufacturer instructions (Meso Scale Diagnostics, Rockville, Maryland, USA). Briefly, reagents were prepared, including calibrator standards. Plates were washed in washing buffer. For all assays, 50 µL of either sample or calibrator were added to each well followed by incubation for 2 hours and 3 washing steps. Samples were then incubated in detection antibody for 2 hours followed by 3 washing steps and addition of 2X Read Buffer to each well. Data were expressed as mean +/− standard error mean (**Supplementary Figures 1 and 2**). Six lists of dysregulated protein levels were generated: #1: Angiogenic and inflammatory factors dysregulated in the blood of mothers who gave birth to a female, #2: Angiogenic and inflammatory factors dysregulated in the blood of mothers who gave birth to a male, #3: Angiogenic and inflammatory factors dysregulated in the umbilical cord blood of females, #4: Angiogenic and inflammatory factors dysregulated in the umbilical cord blood of males, #5: Compilation of angiogenic and inflammatory factors dysregulated in the blood of mothers who gave birth to a female with factors dysregulated in the umbilical cord blood of female neonates, and #6: Compilation of angiogenic and inflammatory factors dysregulated in the blood of mothers who gave birth to a male with factors dysregulated in the umbilical cord blood of male neonates.

### STRING analysis

The two compiled lists (#5, #6) were submitted to a *STRING* analysis to research significantly enriched functional protein-protein interactions (https://string-db.org/; version 11.5). The analysis consisted of identifying predicted associations for a group of proteins based on bioinformatic sources including text mining, databases, experiments, co-expression, neighborhood, gene fusion and co-occurrence (33). The outputs are *i*) a graphical network of predicted associations, displaying proteins as nodes and interactions as edges. Confidence of PPI is visualized by line thickness corresponding to interaction scores: from the thinnest to the thickest lines, low (0.15), medium (0.4), high (0.7) and highest (0.9) confidence, *ii)* a statistical analysis which provides the average node degree (reflecting the number of interactions of proteins within the network) and the PPI enrichment p-value (indicating that connections are not random). Such an enrichment indicated that proteins were at least partially biologically connected as a group.

### ShinyGO analysis

In order to determine if the enriched protein-protein interactions raised by the *STRING* analysis could belong to signaling pathways, the same set of dysregulated proteins was submitted to a *ShinyGO* analysis (https://bioinformatics.sdstate.edu/go/). The outputs are *i)* Functional names of significantly enriched KEGG pathways and *ii)* for each KEGG pathway, the reference number, the enrichment False Discovery Rate (FDR), the number of genes from the submitted list of proteins belonging to the KEGG pathway, the total number of genes of the KEGG pathway and the fold enrichment.

### Statistical analysis

Statistical analysis of the predicted protein–protein interactions (PPIs) was conducted using the *STRING* Network Analysis database (https://string-db.org). Statistical significance was assessed via the PPI enrichment p-value, which compares the number of observed network edges to the expected number for a random protein set. p-values were adjusted for multiple comparisons using the Benjamini–Hochberg correction. Statistical analysis of enriched biological pathways was performed using *ShinyGO* analysis (https://bioinformatics.sdstate.edu/go/). Statistical significance was calculated by using the −log10 (FDR; false discovery rate) method. P-value is derived from the hypergeometric distribution. A significant FDR cutoff was fixed at 0.05. Statistical analysis of relative protein levels was performed using Prism (version 8.0.2; GraphPad Software, La Jolla, CA, USA). Normality was assessed using the Shapiro-Wilk test. Comparison between non-consuming (non-exposed) and alcohol-consuming pregnant women (*in utero* exposed to alcohol) were performed using unpaired t-test for normally distributed continuous variables, Mann-Whitney *U* test for non-normal distributed continuous variables and Fisher’s exact test for categorial variables. Statistical significance was set at p<0.05 with a trend defined as 0.05<p<0.1 (34).

### Study approval

The cohort consisted in 31 individuals in the Control group (no alcohol consumption) and in 28 women in the Alcohol group (alcohol consumption during pregnancy). Sample collection was performed under the protocol n°2016-A01563-48. All participants provided written informed consent for the study.

## Supporting information

Supplementary Table 1 without exact dates

## Data availability

Raw data are available in the supporting data table 1.

## Author contributions

GBJ and MS designed the study. SC and LC carried out experiments. BH, BV, DAL, GP, LA, MF, MJB, PCG and VE provided resources. SC, GBJ and MS performed analysis. SC, GBJ and MS wrote the manuscript. GBJ and MS acquired funding.

## Acknowledgements

We thank Bruchbach V, Marty N, Caillot F from the Department of Clinical Research and Innovation, Rouen University Hospital; Bubenheim M from the Department of Biostatistics, Rouen University Hospital; Mestre N from the Department of Neonatal Pediatrics and Intensive Care, Rouen University Hospital; Bidar F from the Clinical research, Le Havre Hospital; Arokyanadar M from the Department of Neonatal Pediatrics and Intensive Care, Robert Debré Hospital; Brien A, from the Women-Mother-Child Unit, Morvan-Brest University Hospital.

