## Supplementary Figure 1 for "Alcohol consumption during pregnancy dysregulates maternofetal angiogenic and inflammatory factors with sex specificities"

### Angiogenic factors

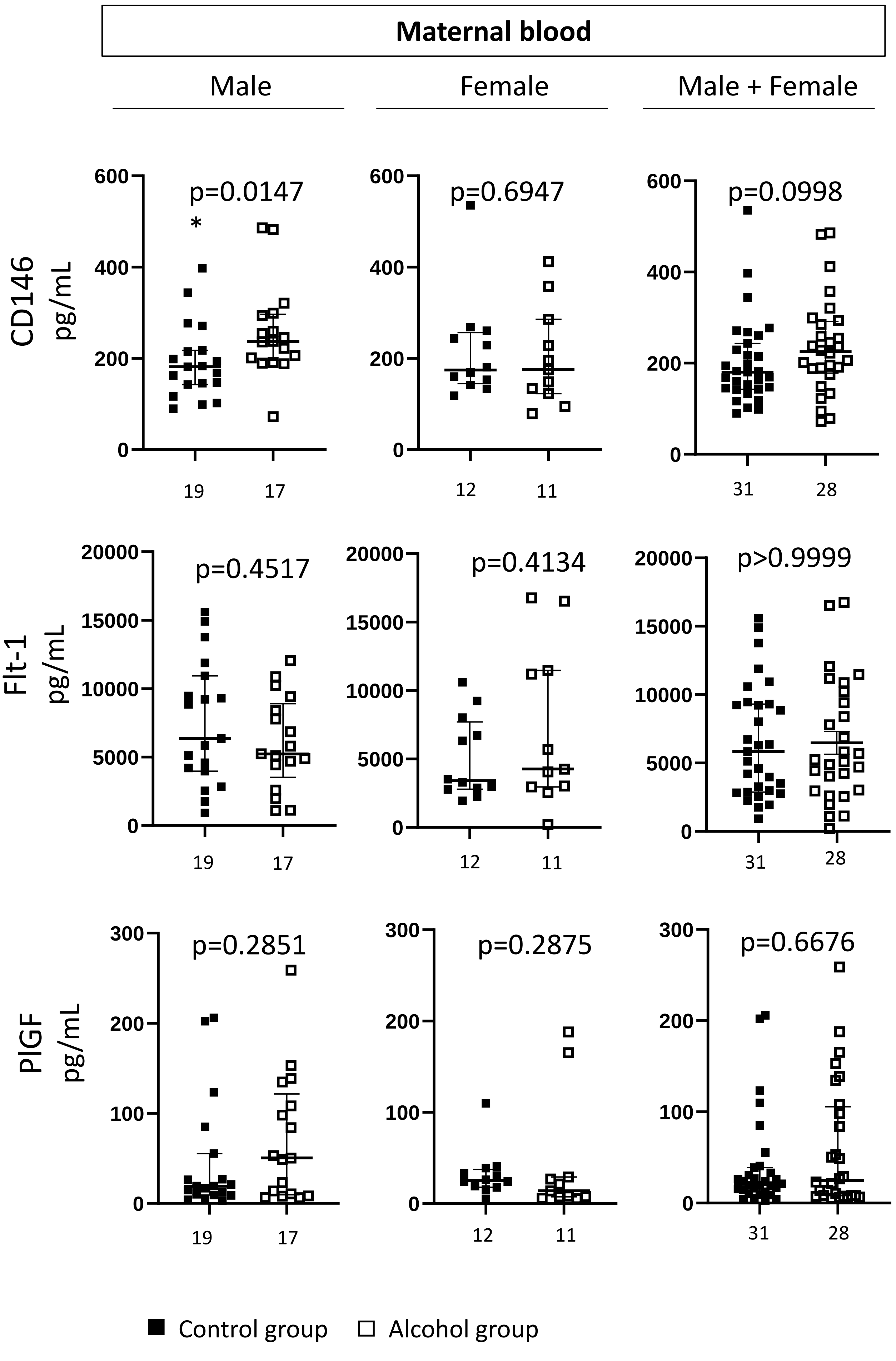

### Angiogenic factors

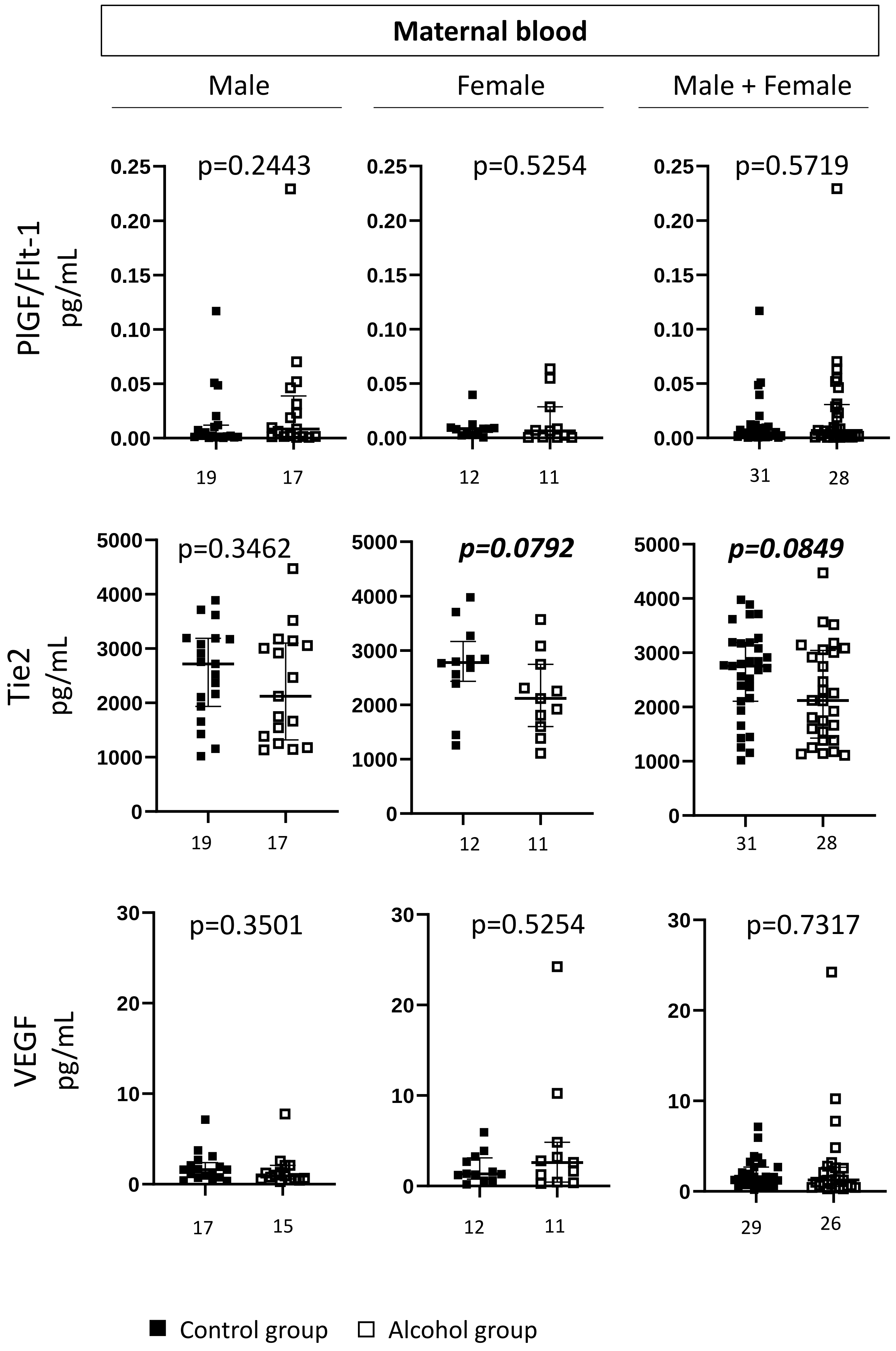

### Angiogenic factors

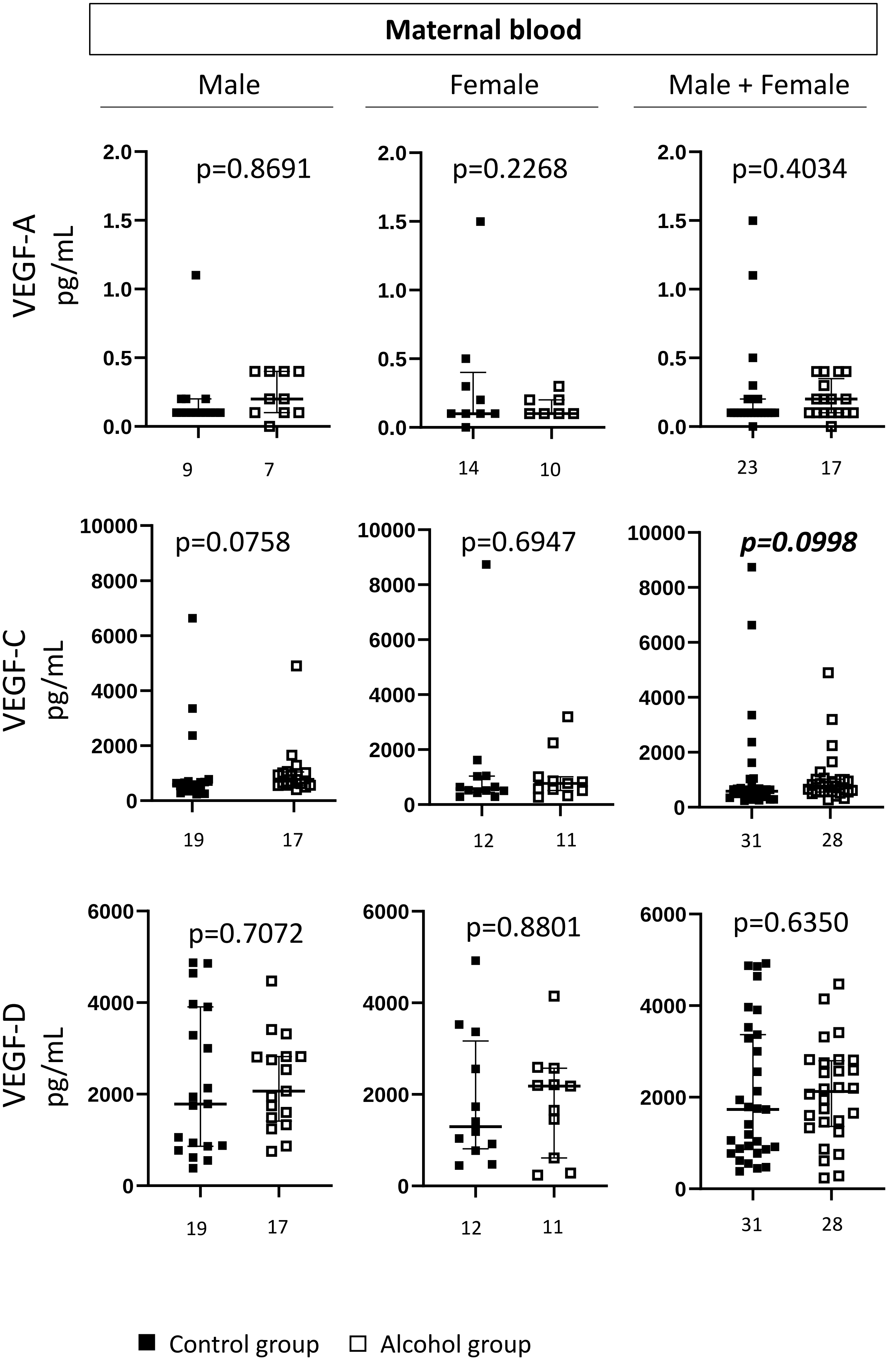

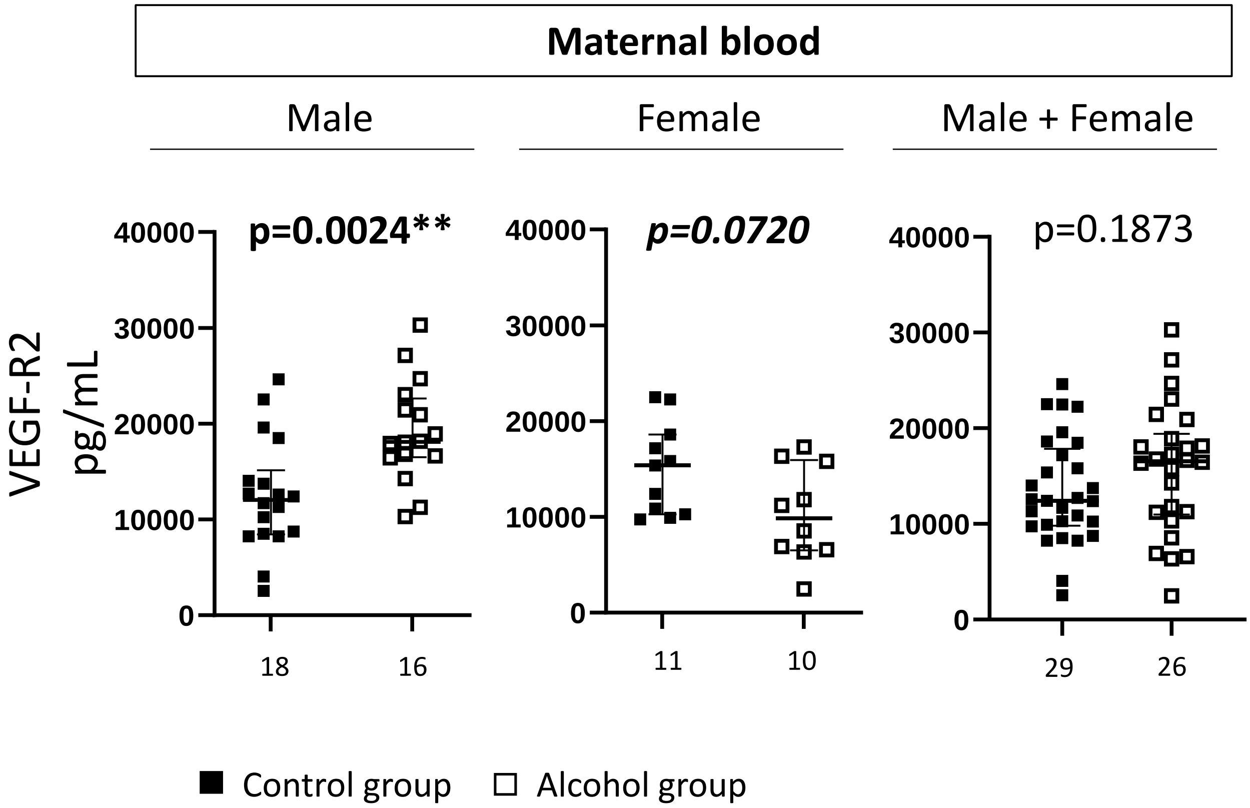

Maternal blood
Male
Female
Male + Female
VEGF-R2 pg/mL
p=0.0024**
p=0.0720
p=0.1873
40000
30000
20000
10000
0
18
16
11
10
29
26
Control group
Alcohol group

### Chemokines/Cytokines

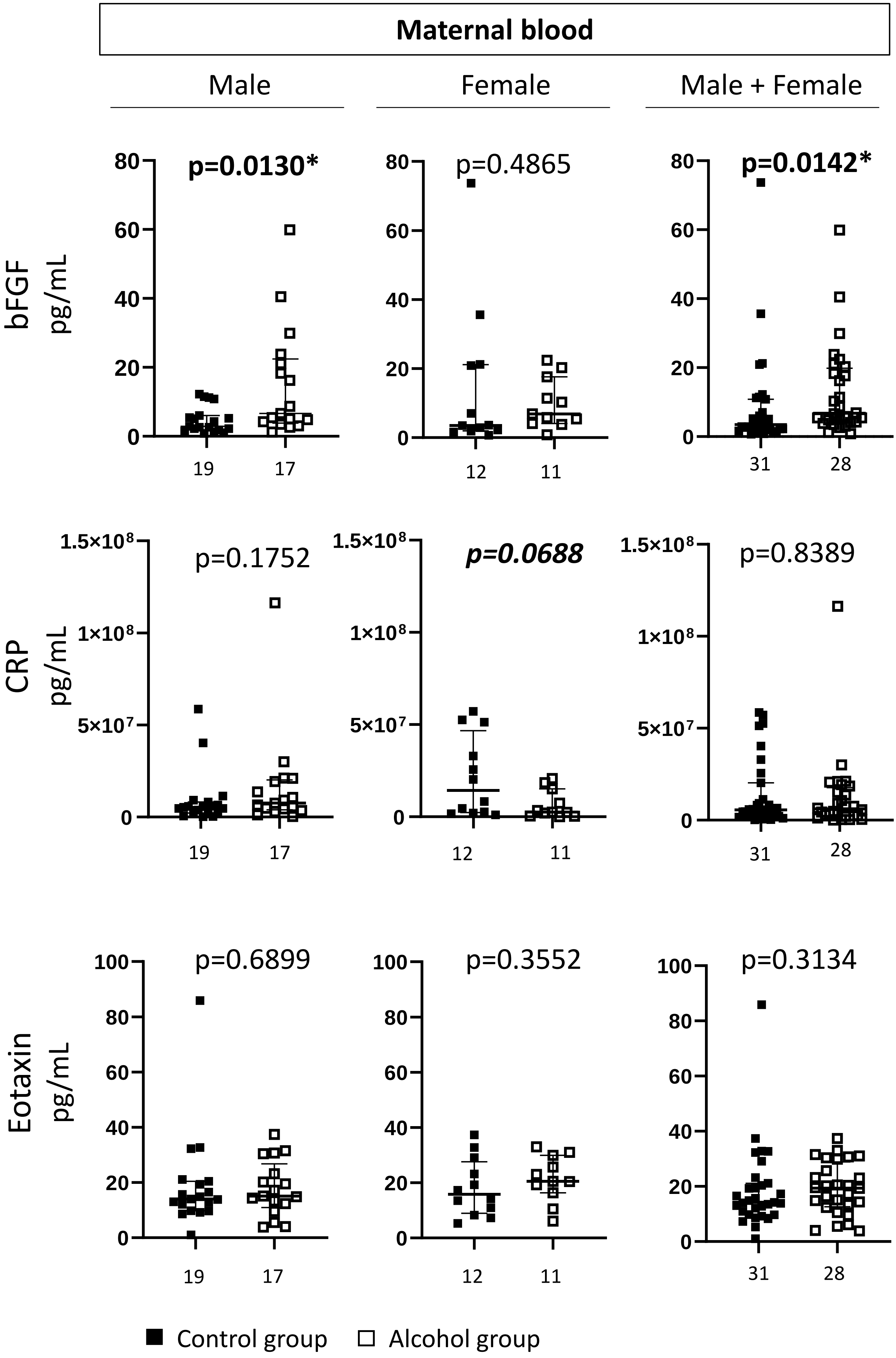

### Chemokines/Cytokines

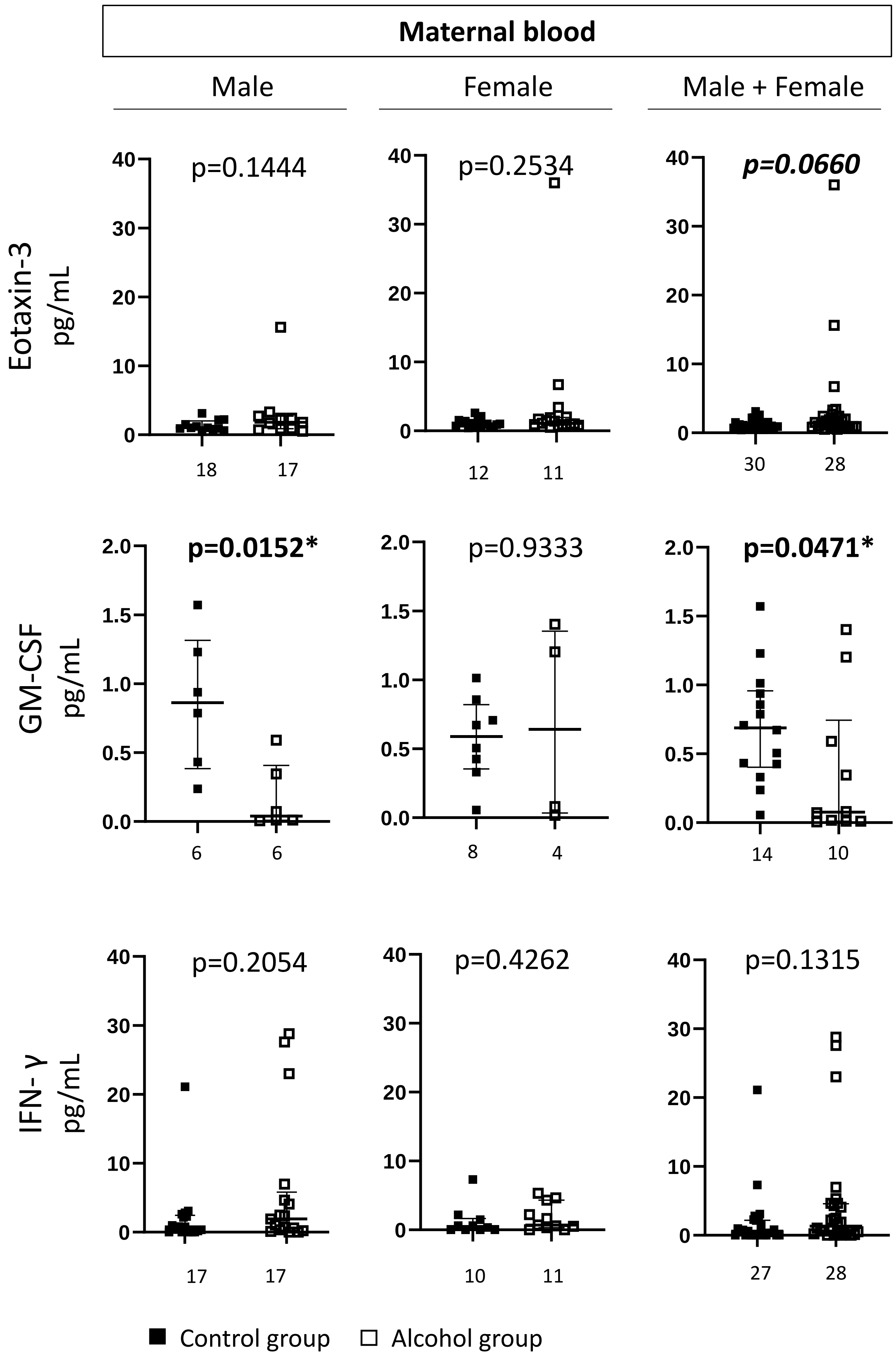

### Chemokines/Cytokines

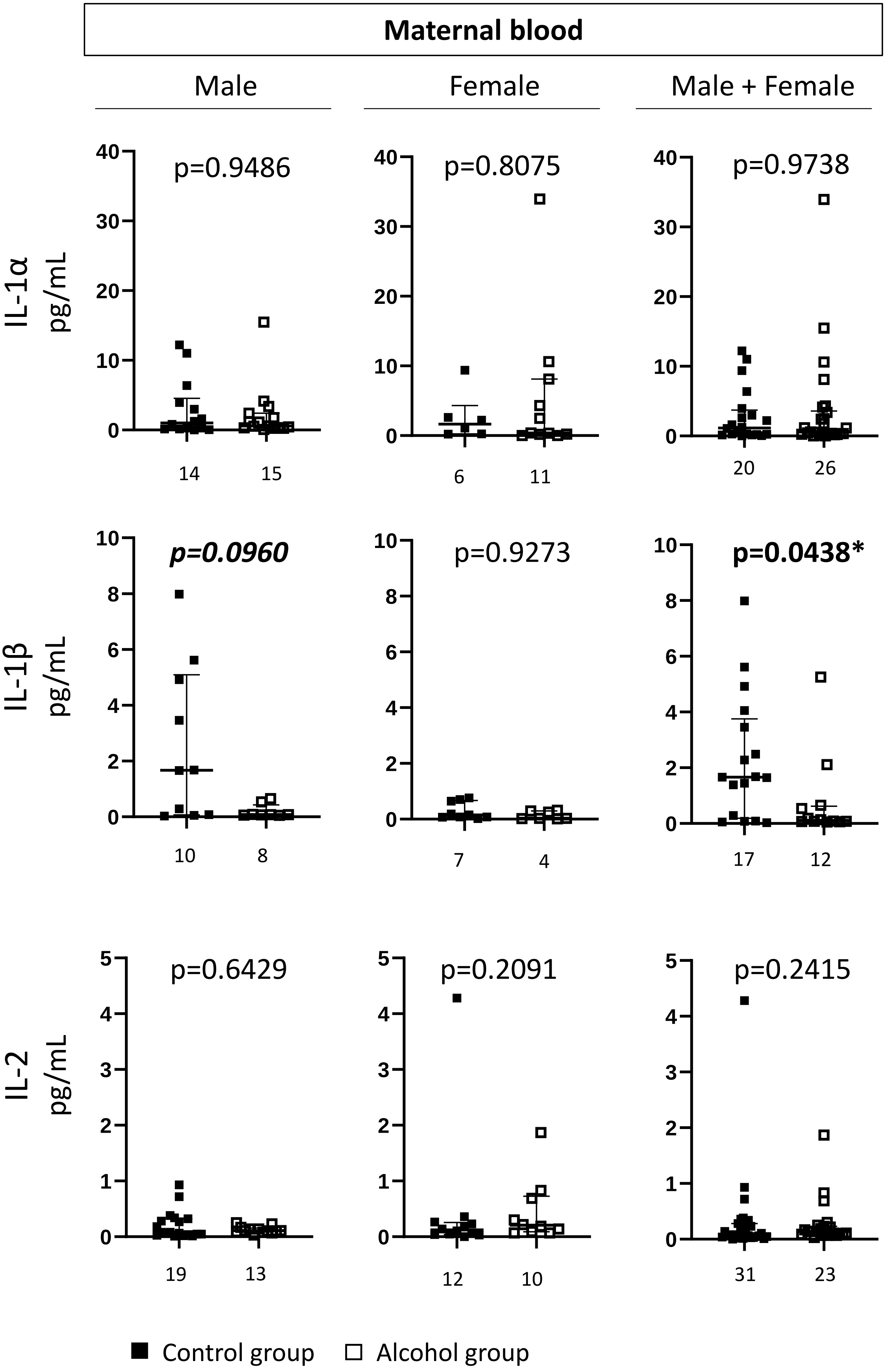

#### Chemokines/Cytokines

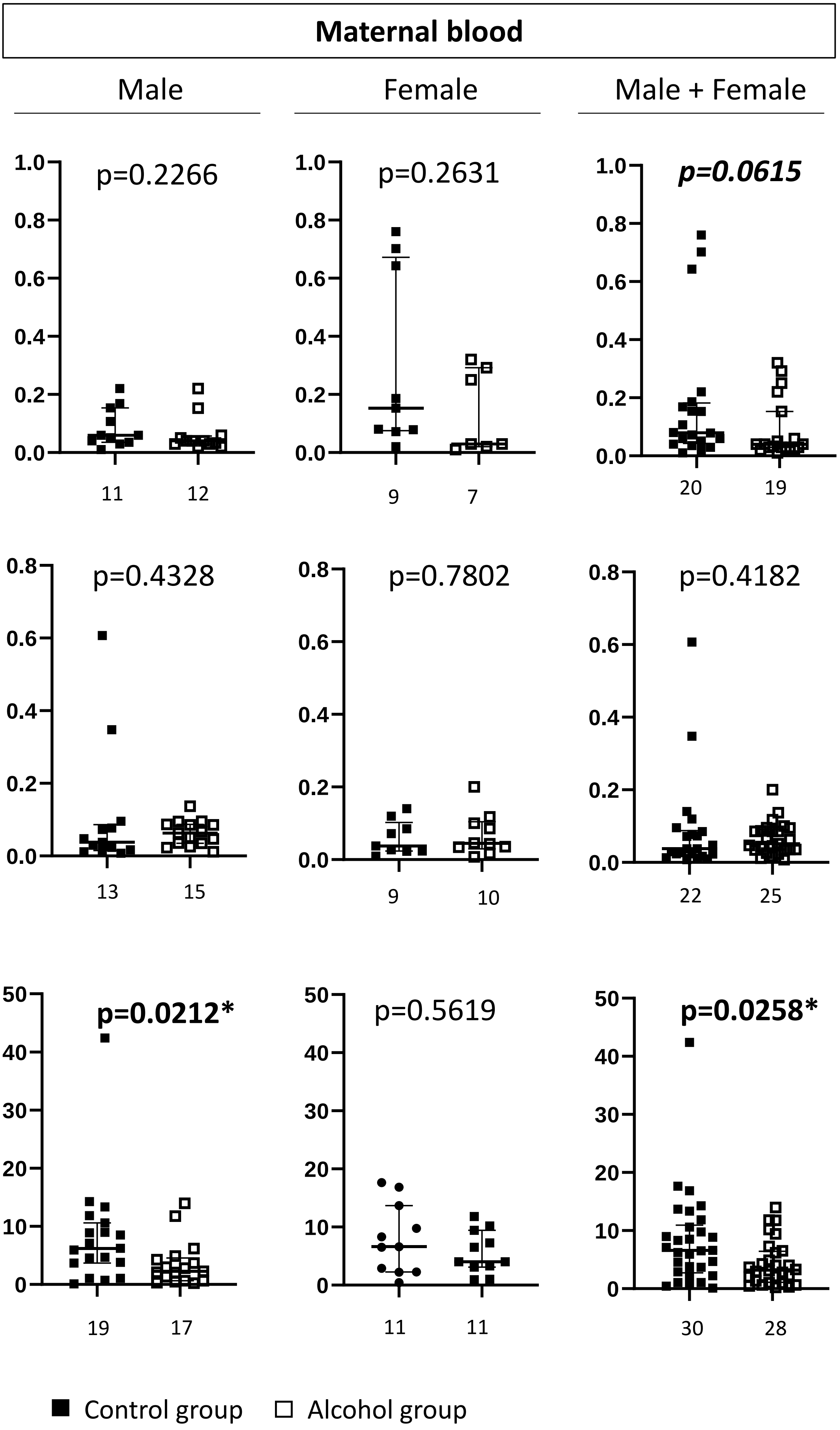

### Chemokines/Cytokines

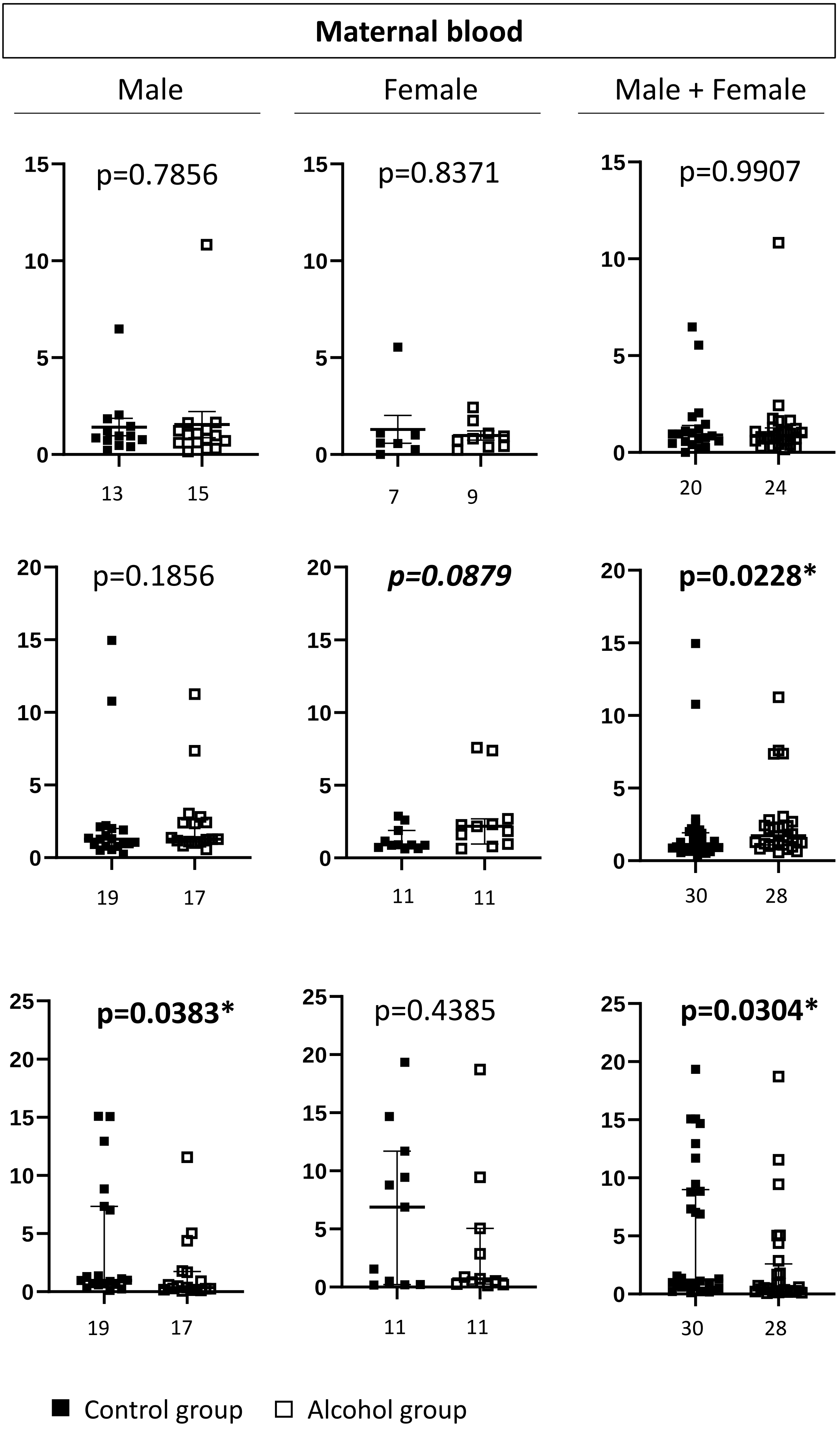

### Chemokines/Cytokines

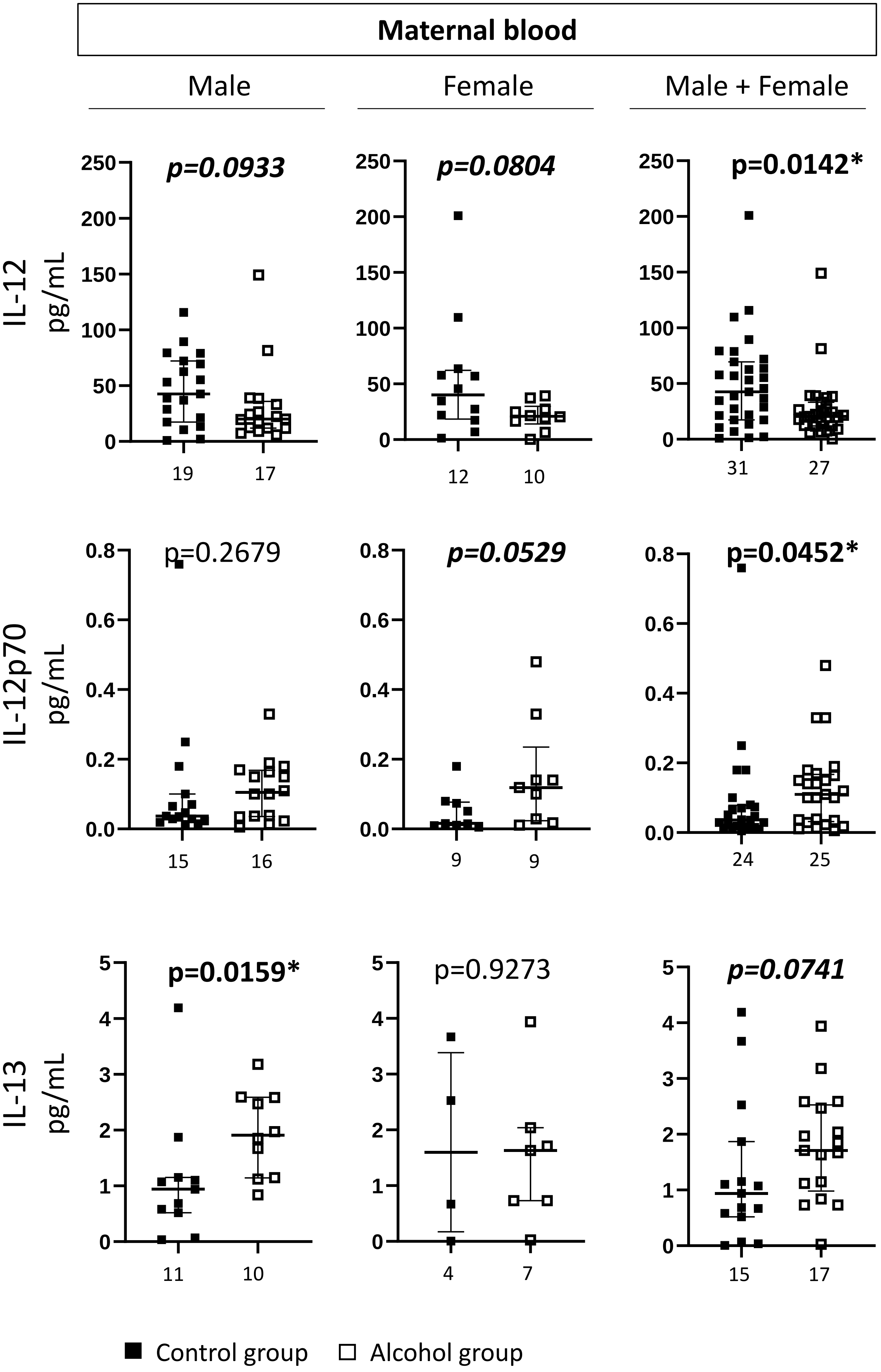

### Chemokines/Cytokines

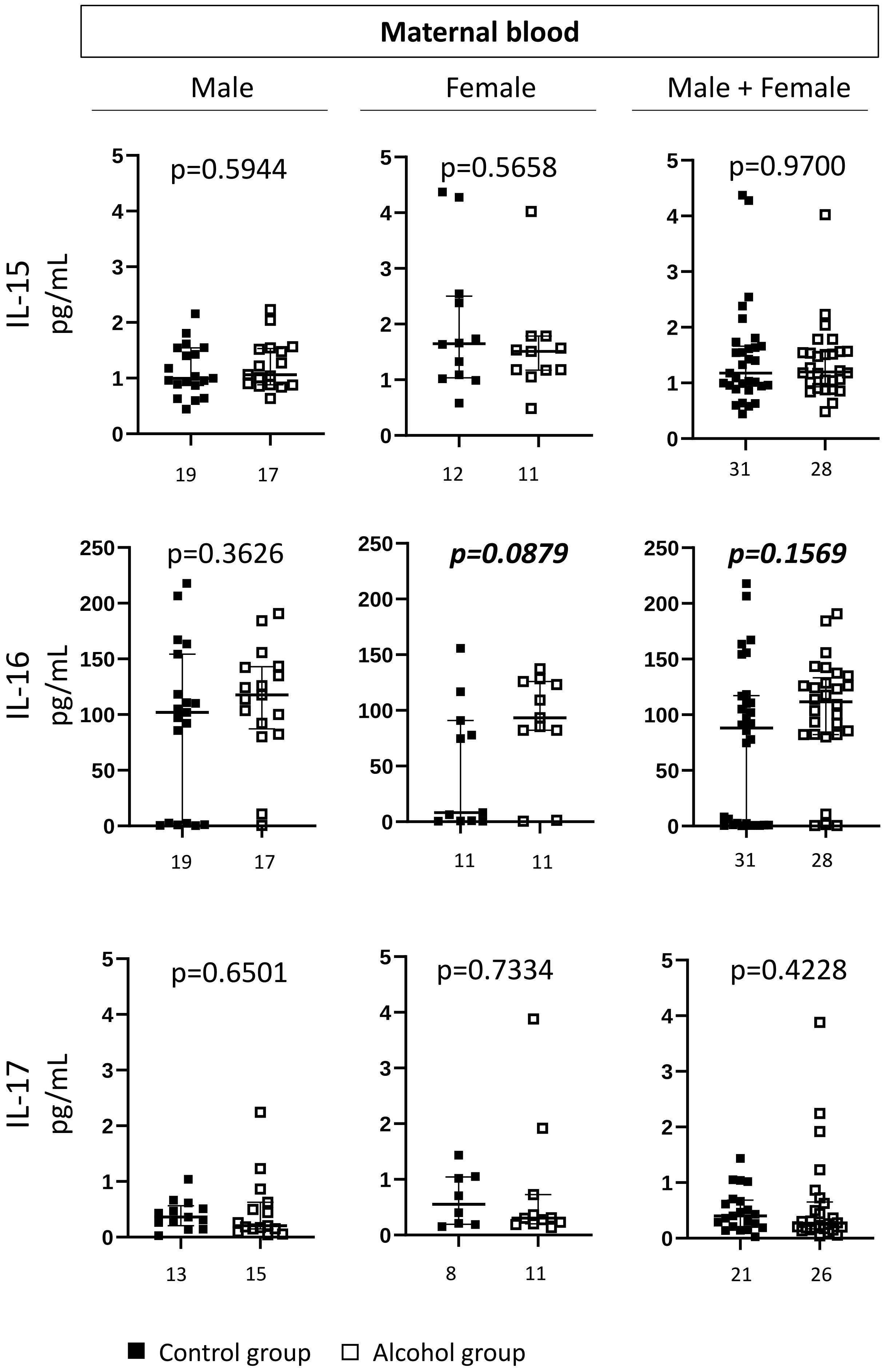

### Chemokines/Cytokines

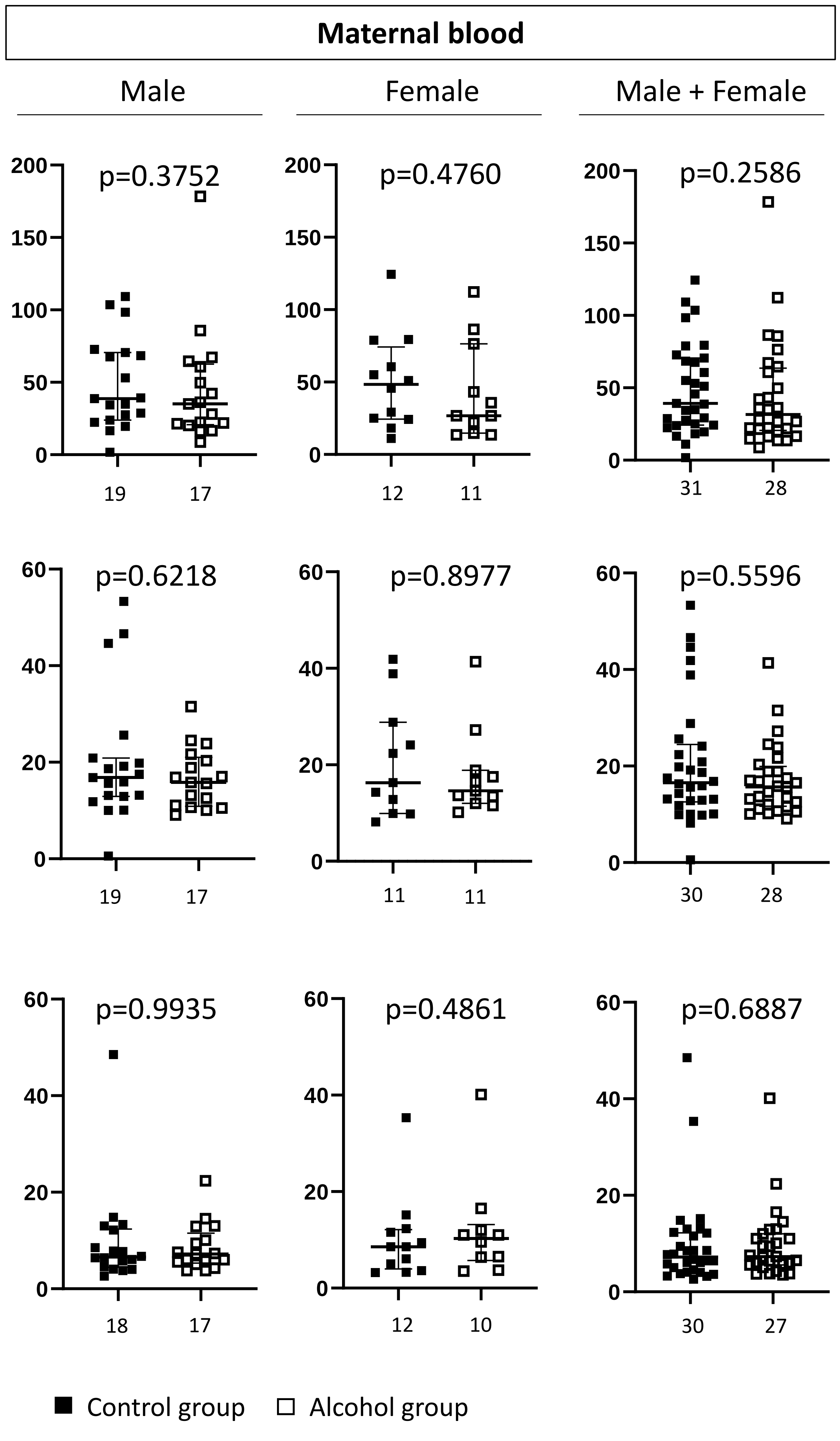

### Chemokines/Cytokines

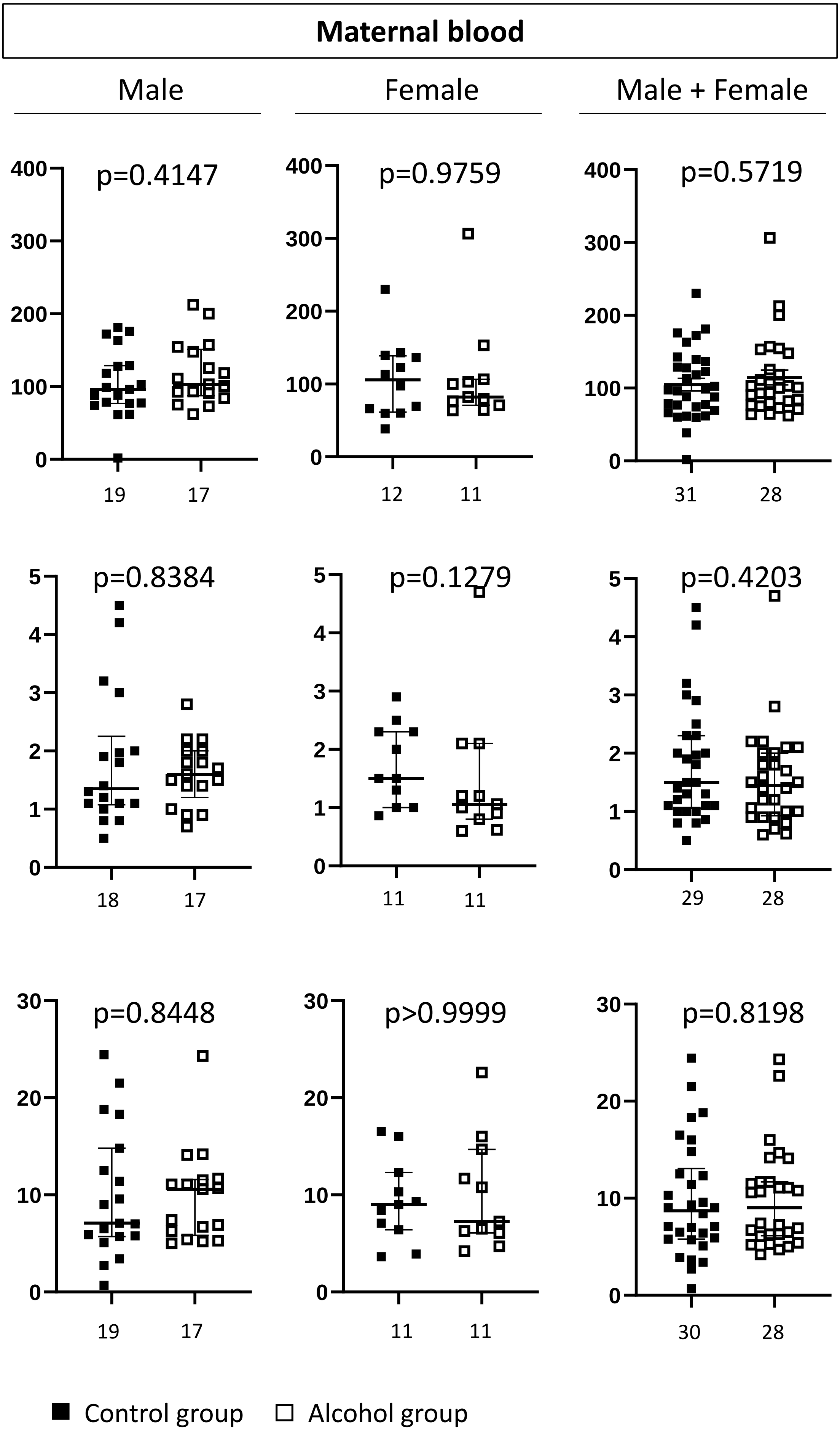

### Chemokines/Cytokines

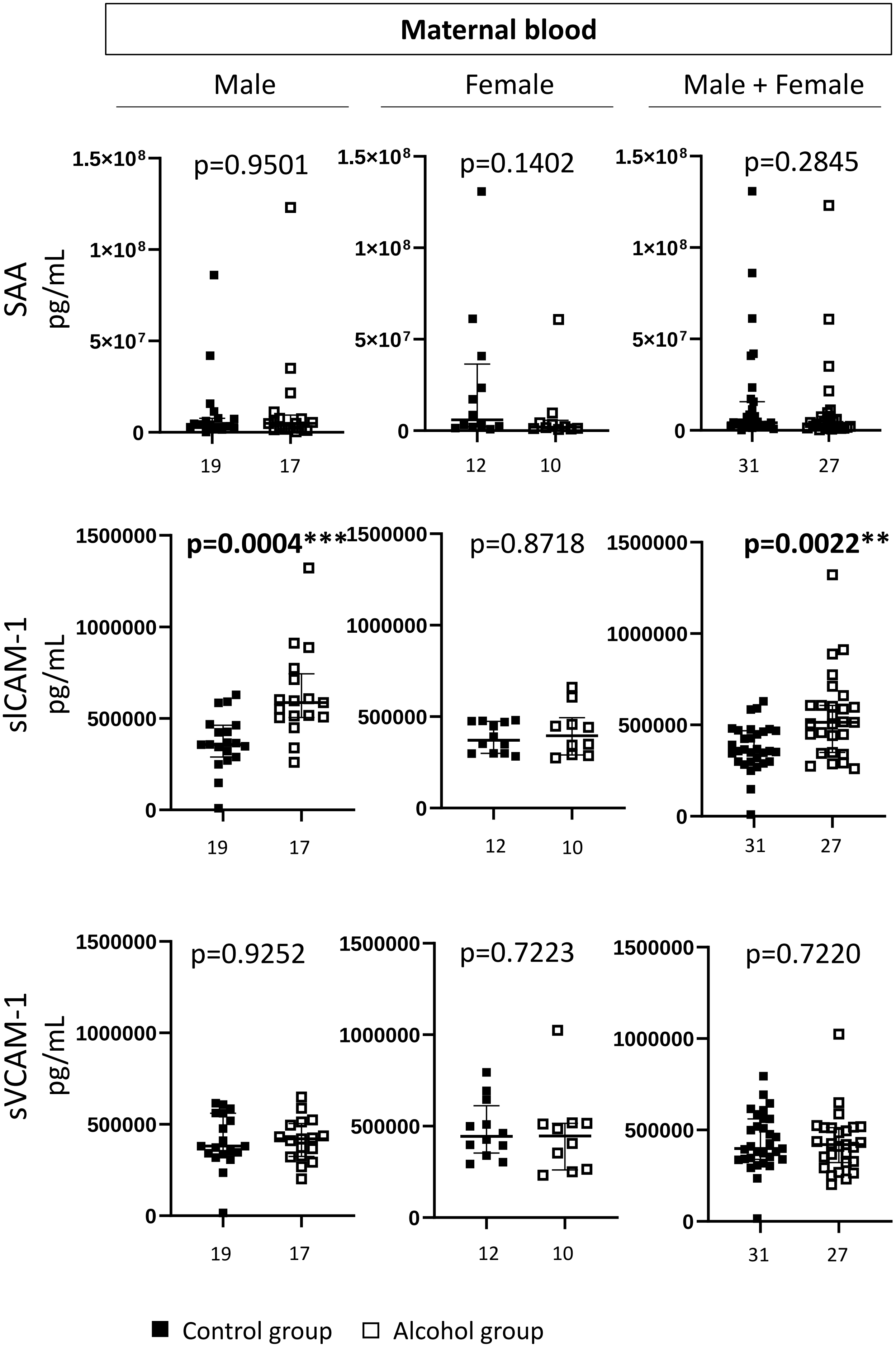

### Chemokines/Cytokines

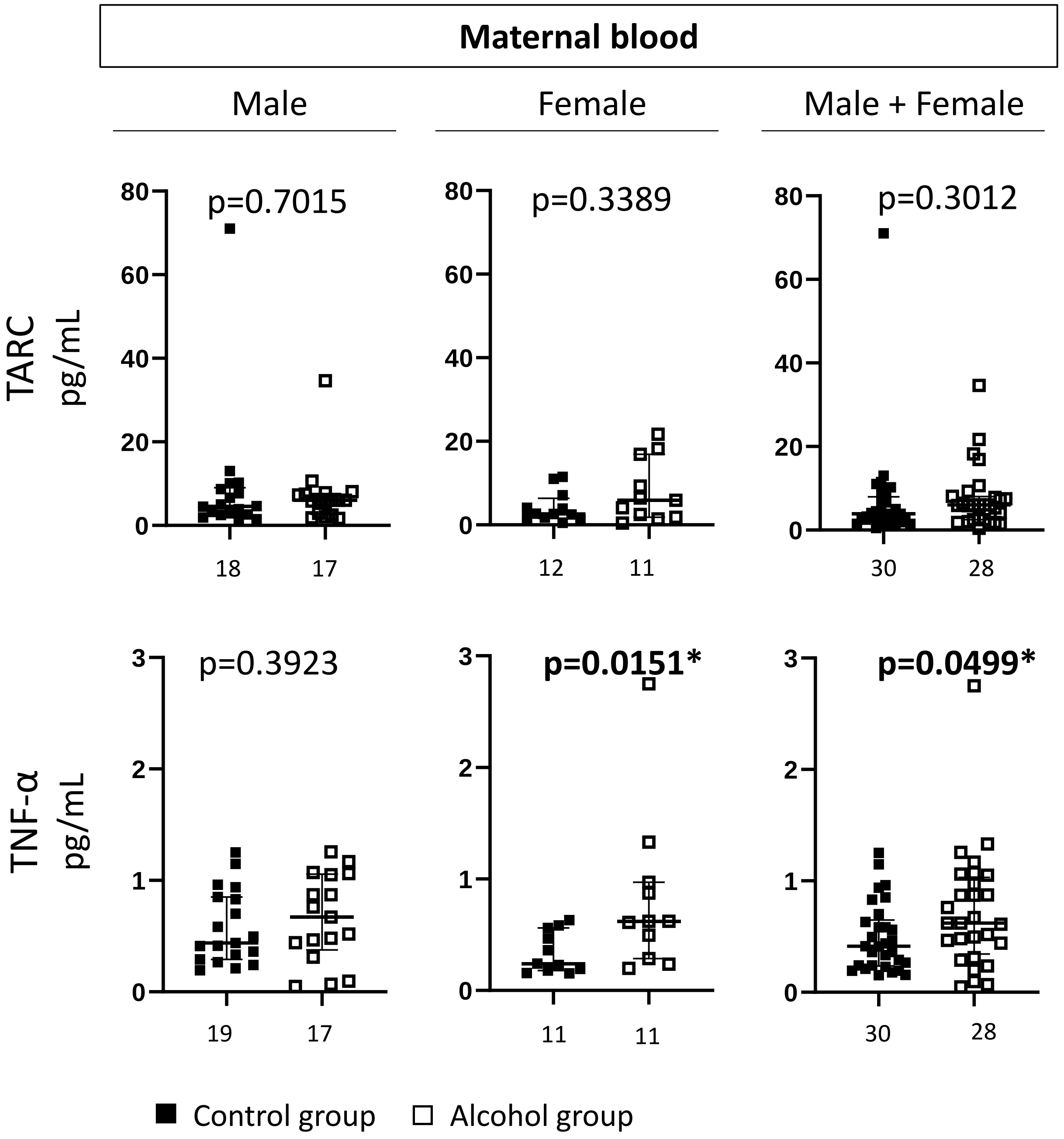


**Supplementary Figure 1. Levels of angiogenic factors, cytokine and chemokine in maternal blood from Control and Alcohol groups.** Numbers in abscissa indicate the number of values (n) per group. Different n values between two factors for a given group, result from missing data. Values from Control and Alcohol groups were compared using the Mann-Whitney test. $p<0.05$ was considered statistically significant (*) and $0.05<p<0.1$ considered as a trend.
