## Supplementary Figure 2 for "Alcohol consumption during pregnancy dysregulates maternofetal angiogenic and inflammatory factors with sex specificities"

### Angiogenic factors

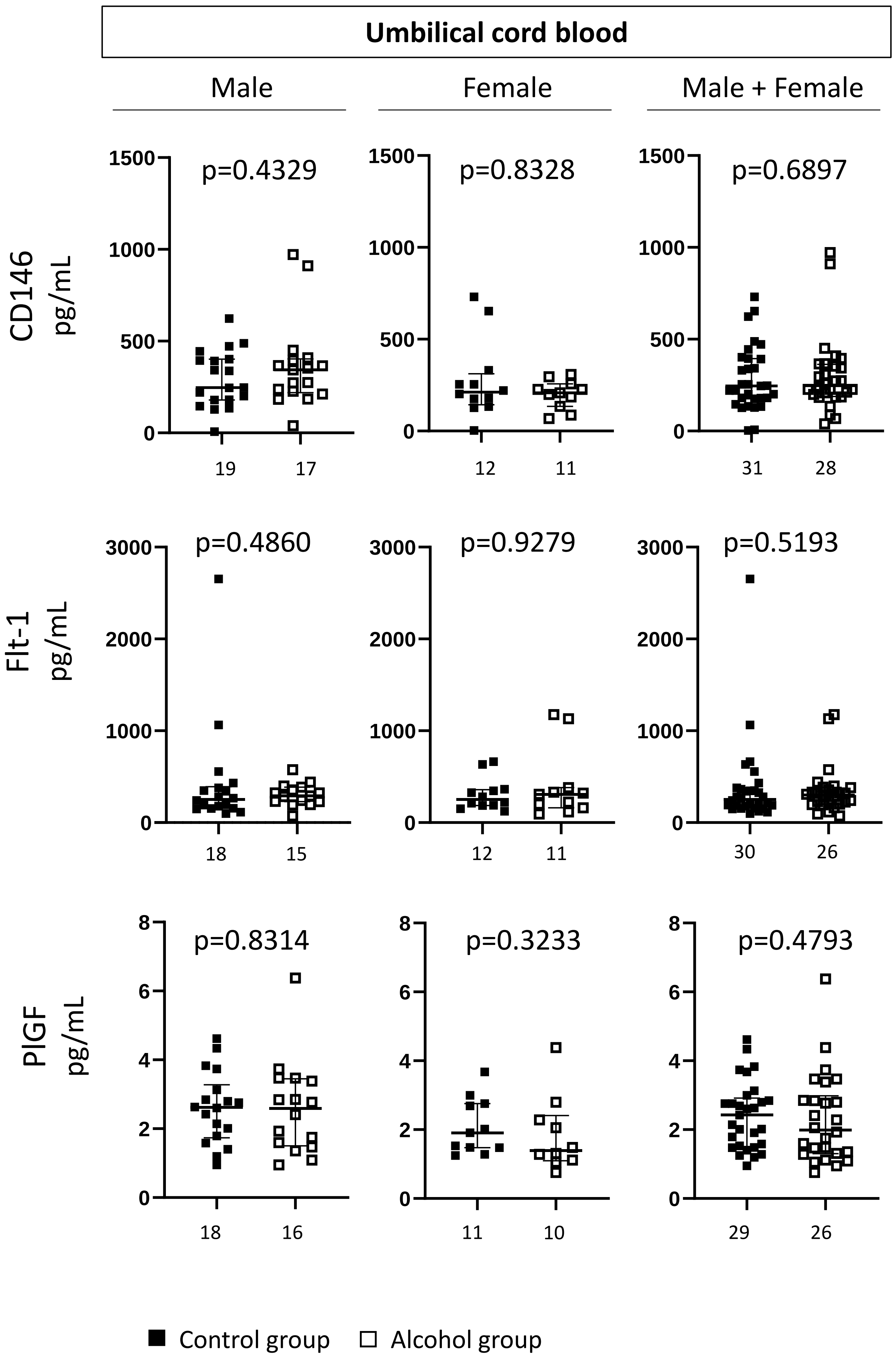

### Angiogenic factors

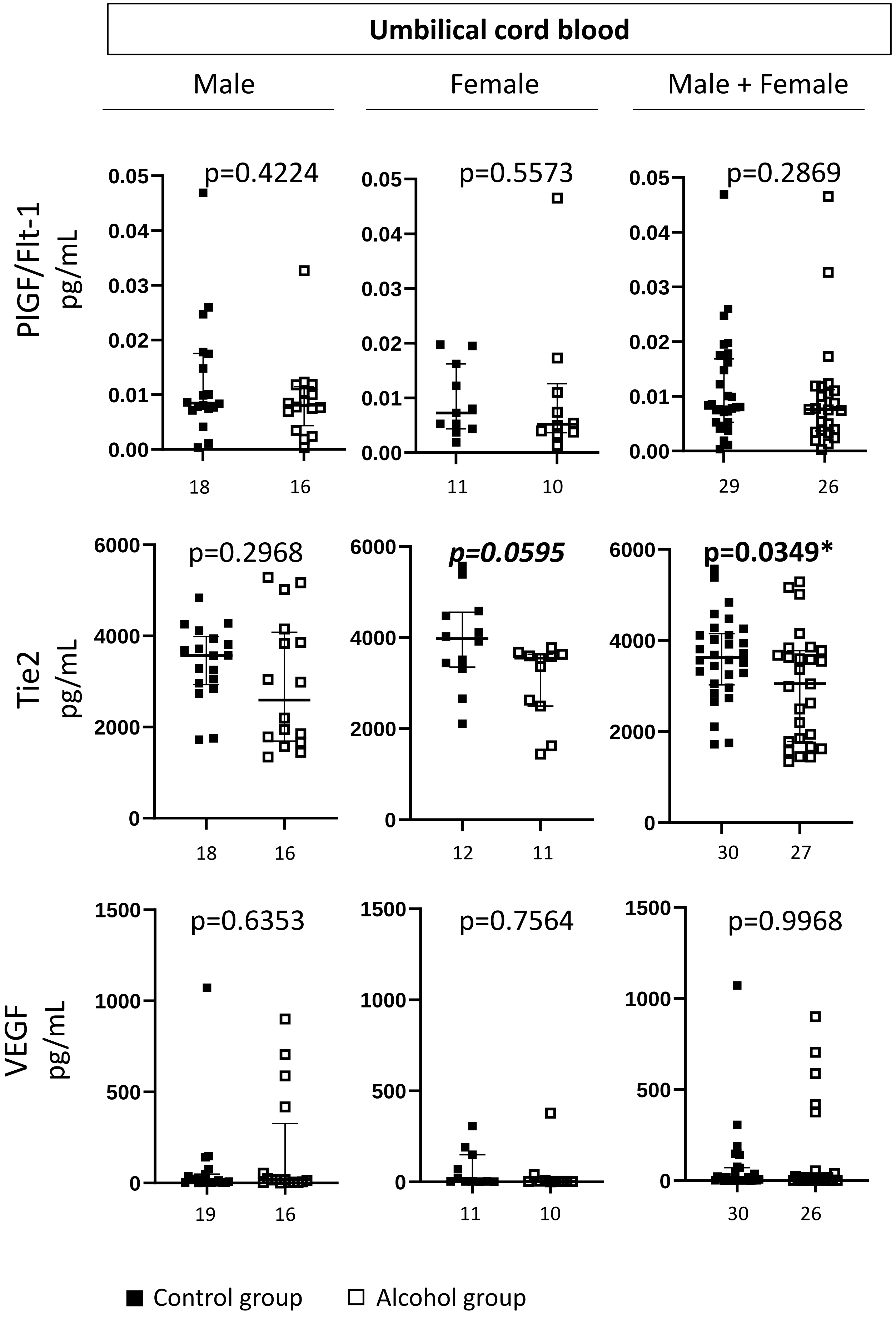

### Angiogenic factors

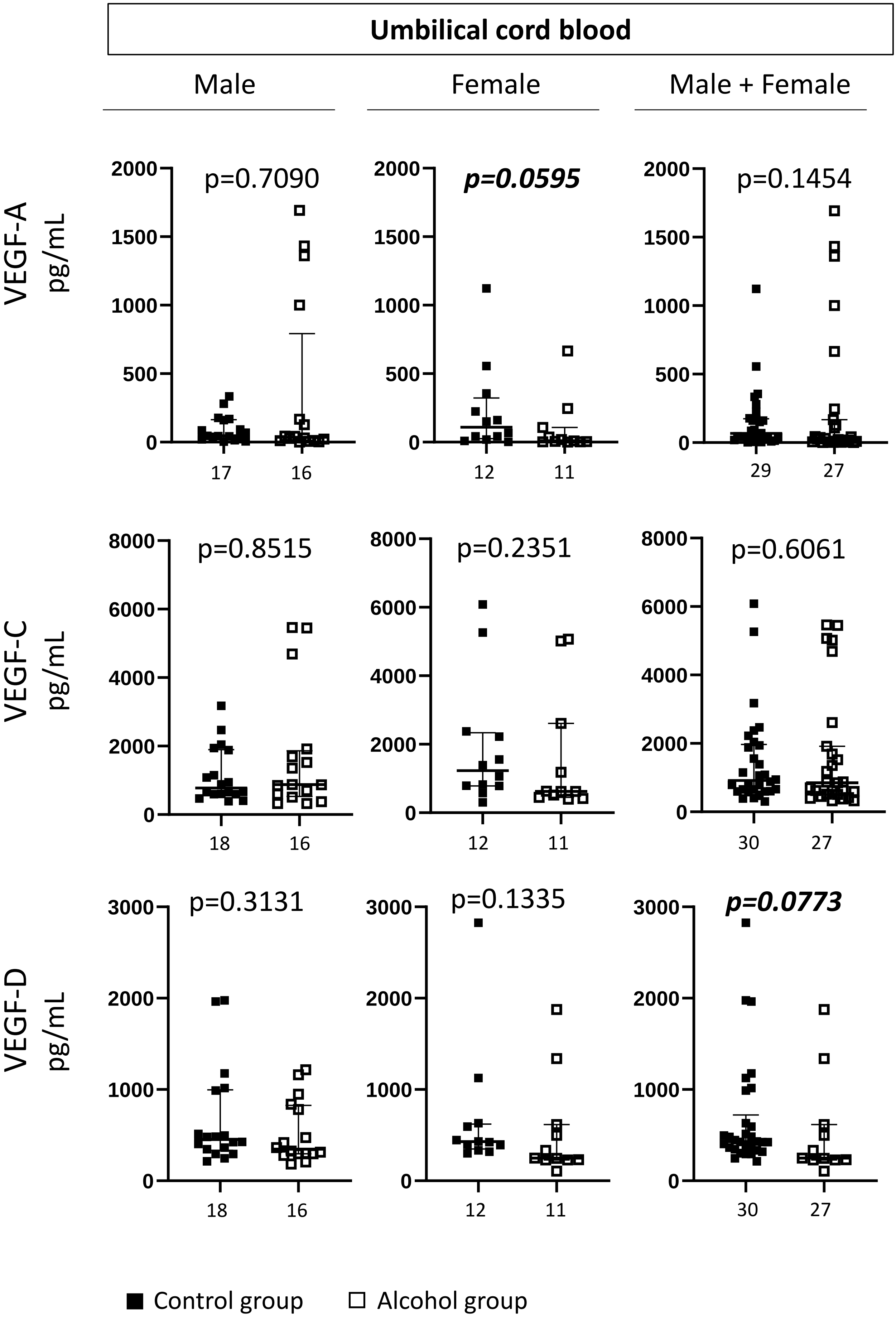

### Angiogenic factors

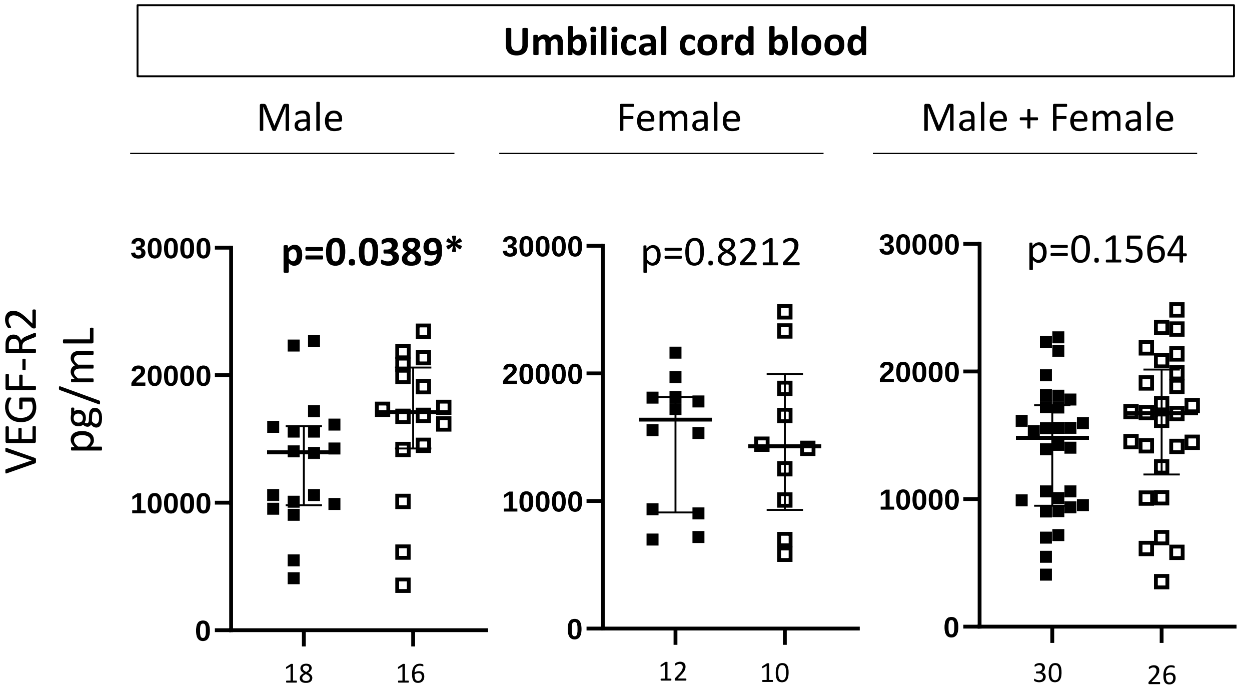


■ Control group ☐ Alcohol group

### Chemokines/cytokines

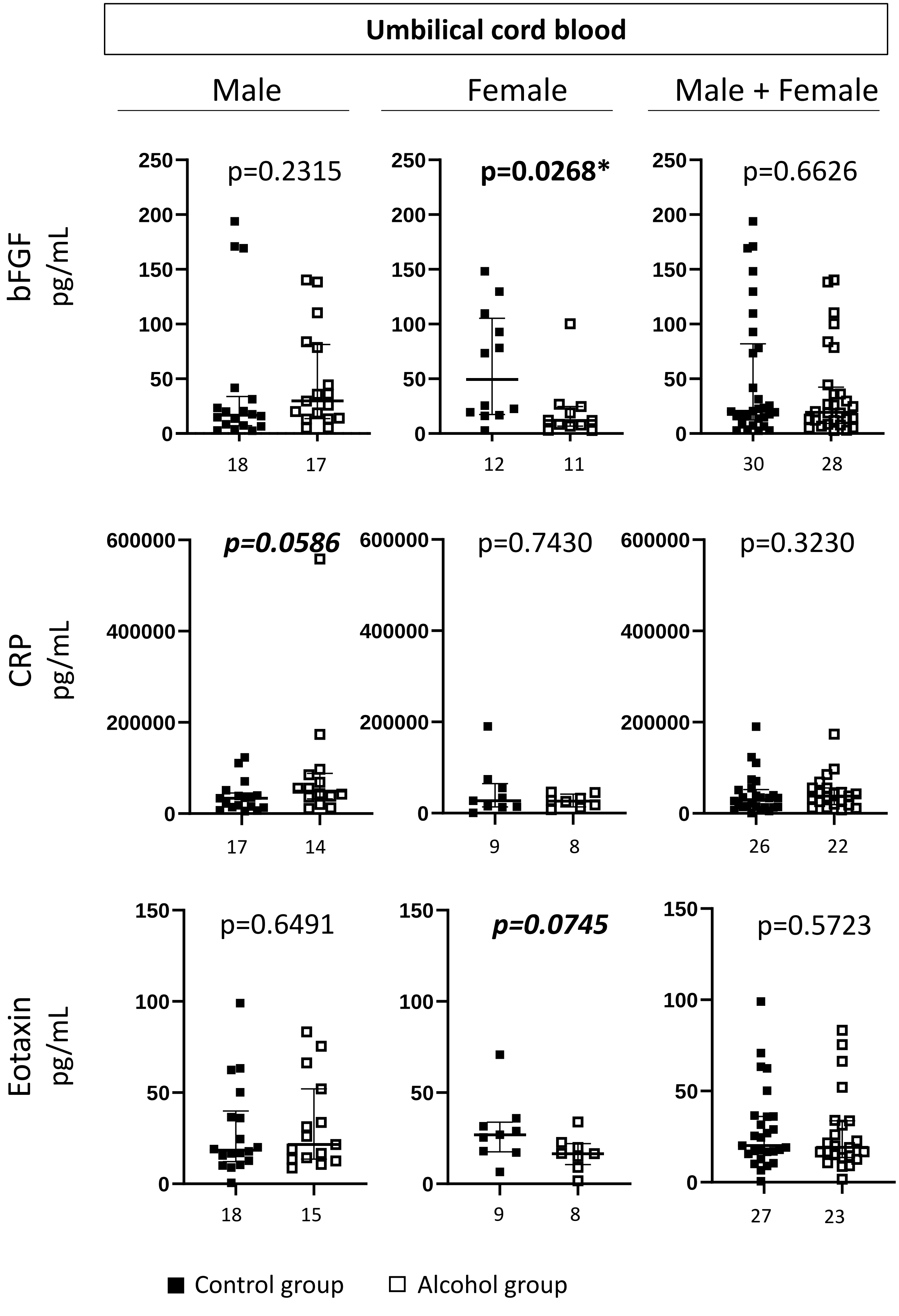

#### Chemokines/cytokines

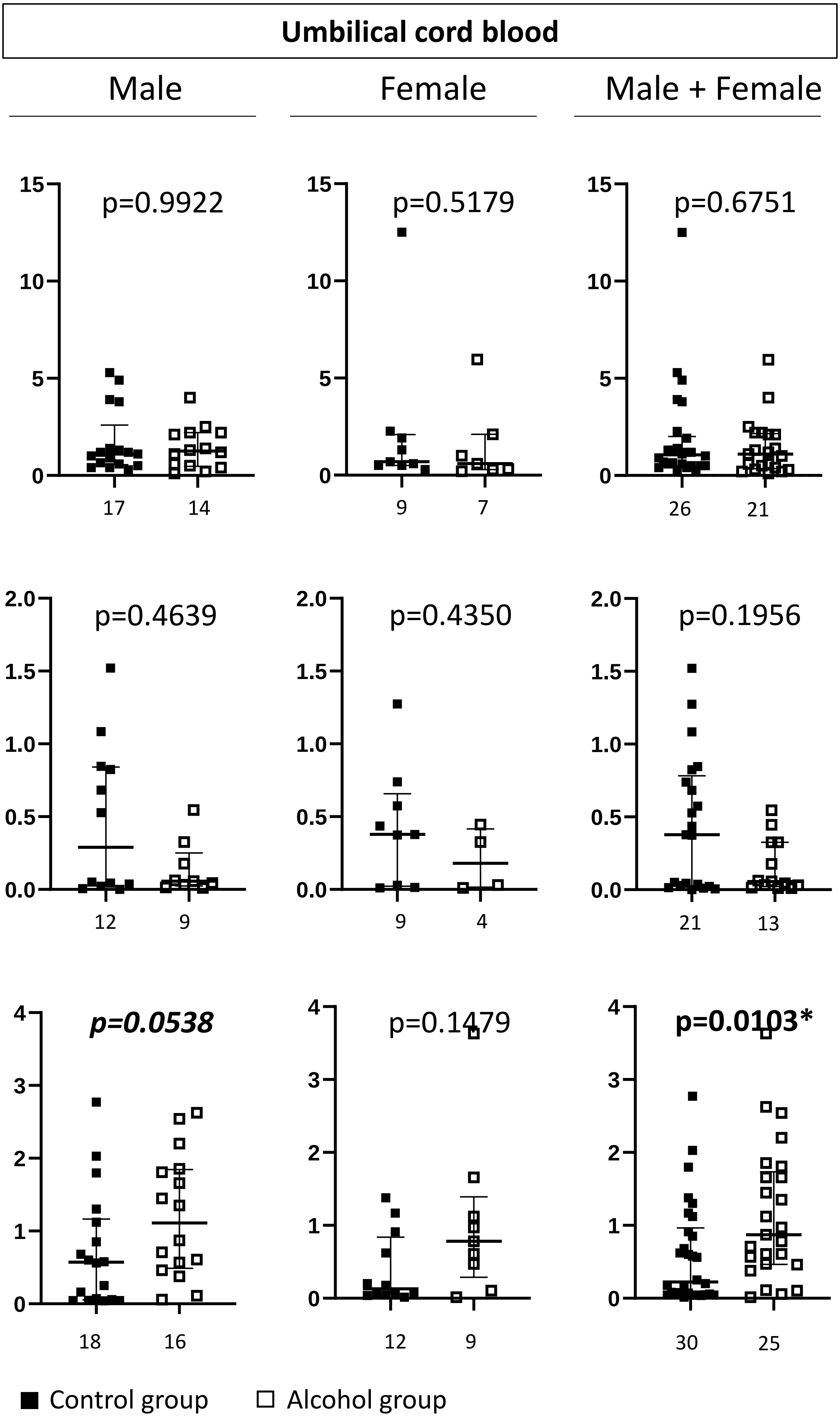

### Chemokines/cytokines

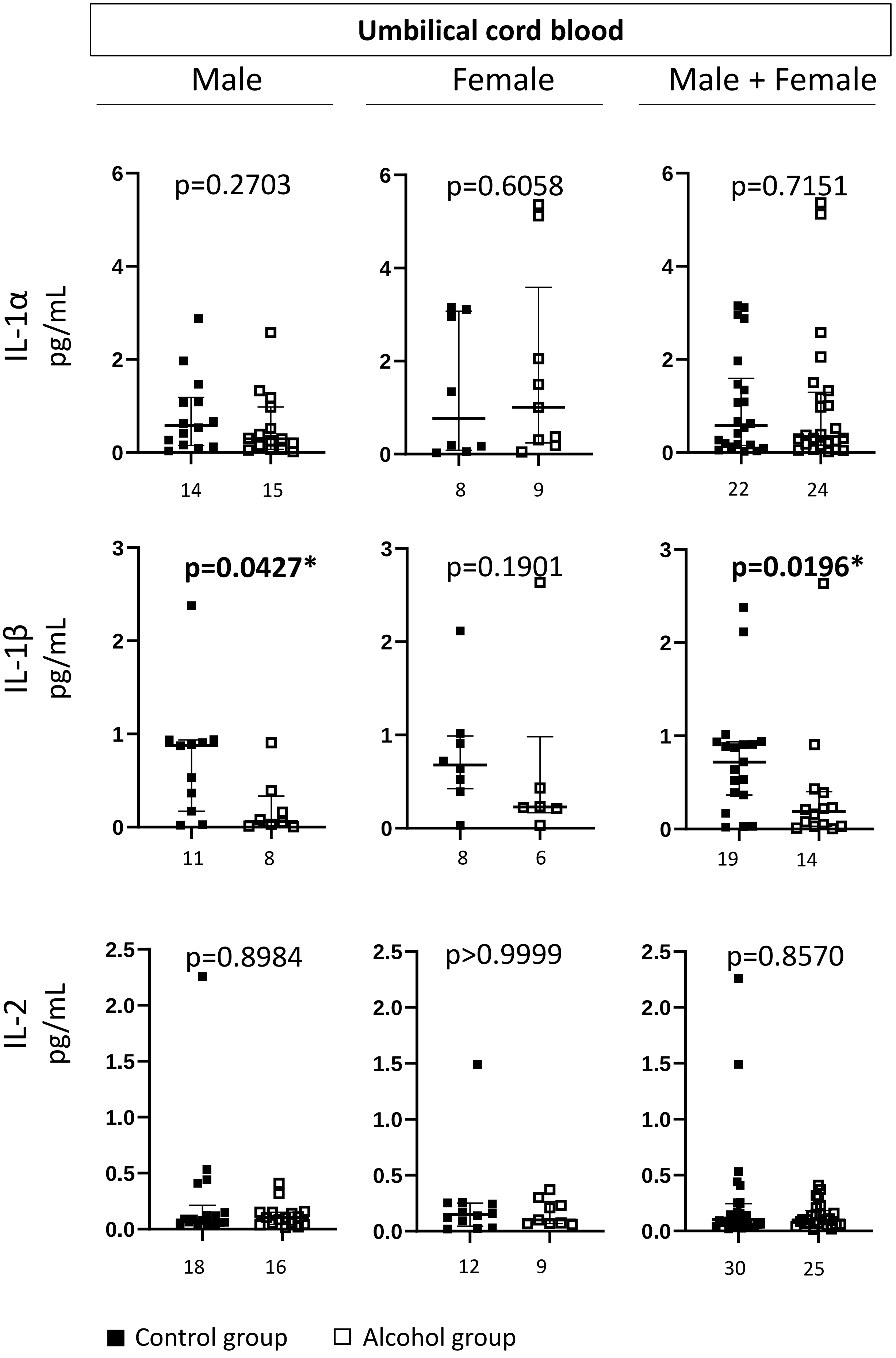

#### Chemokines/cytokines

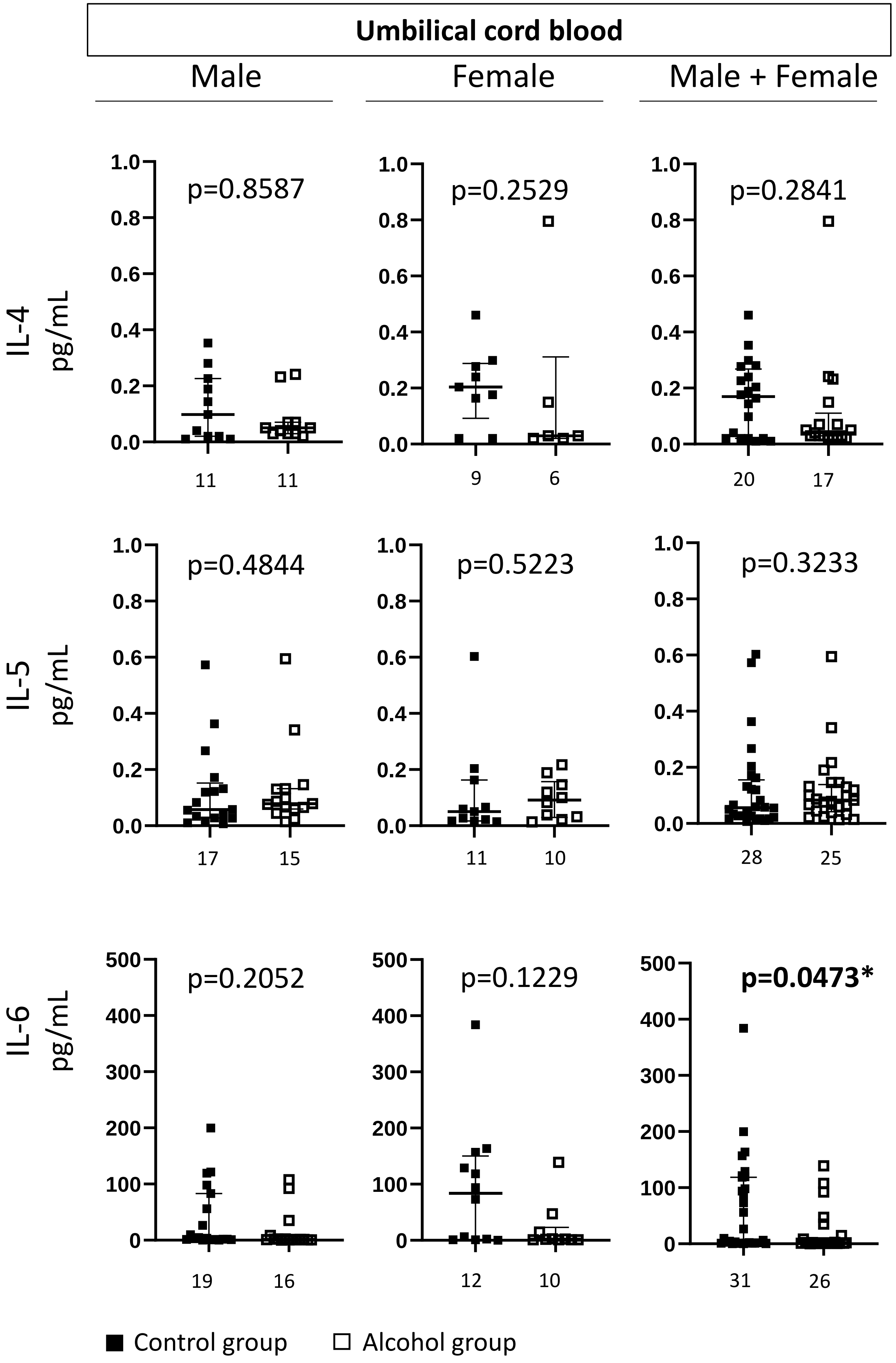

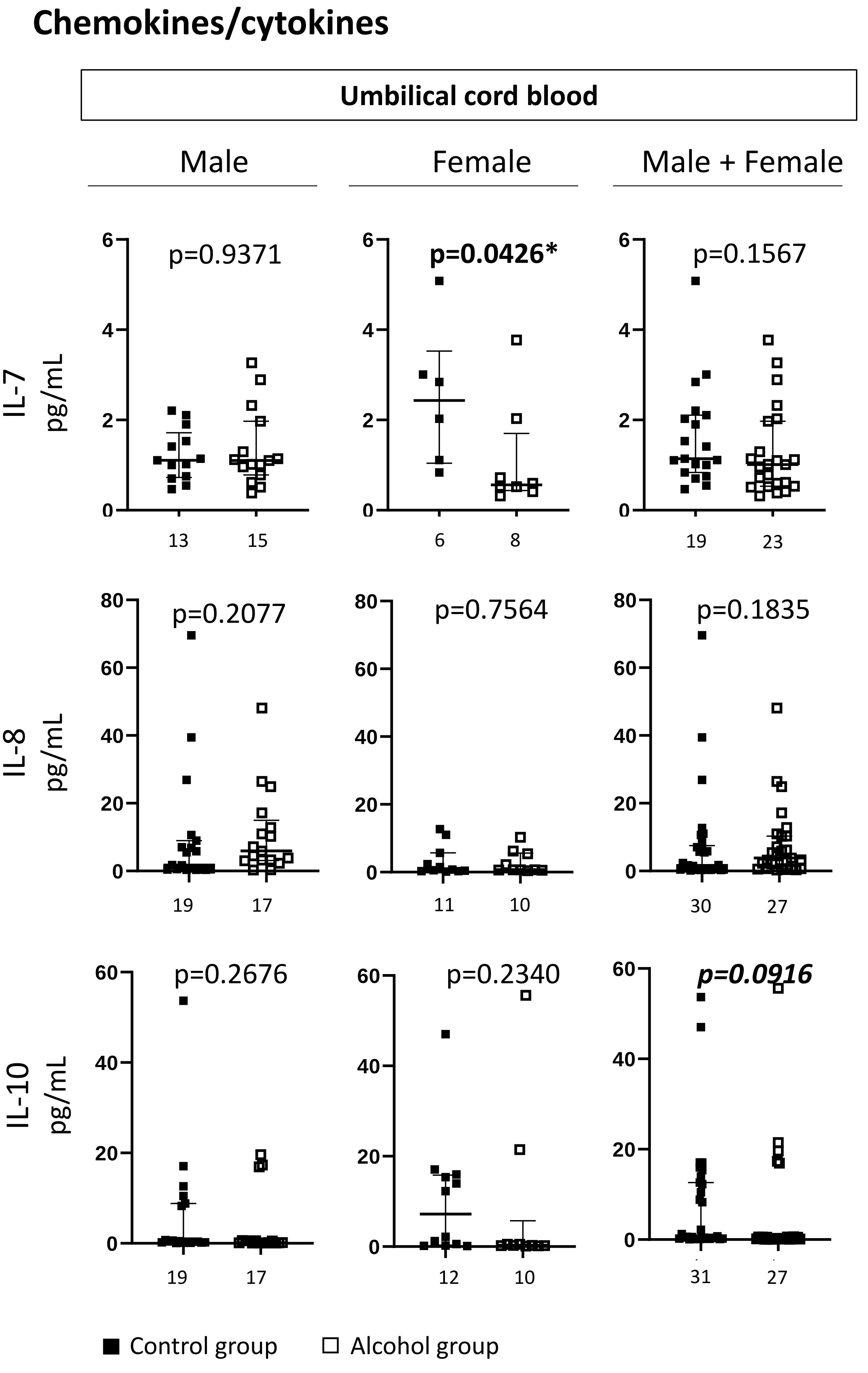

Chemokines/cytokines
Umbilical cord blood
Male
Female
Male + Female
IL-7 pg/mL
p=0.9371
p=0.0426*
p=0.1567
13
15
6
8
19
23
IL-8 pg/mL
p=0.2077
p=0.7564
p=0.1835
19
17
11
10
30
27
IL-10 pg/mL
p=0.2676
p=0.2340
p=0.0916
19
17
12
10
31
27
Control group
Alcohol group

### Chemokines/cytokines

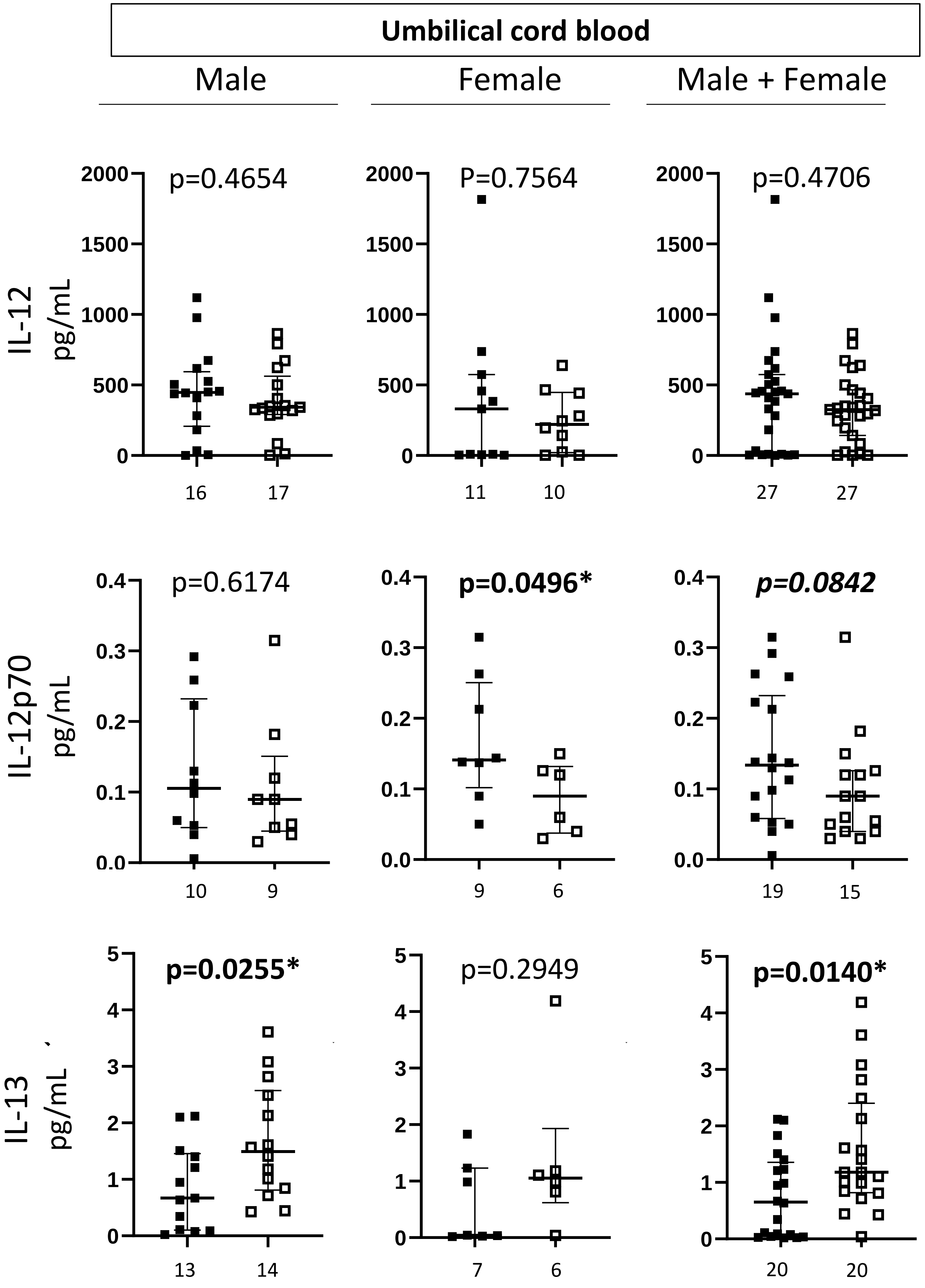


■ Control group □ Alcohol group

#### Chemokines/cytokines

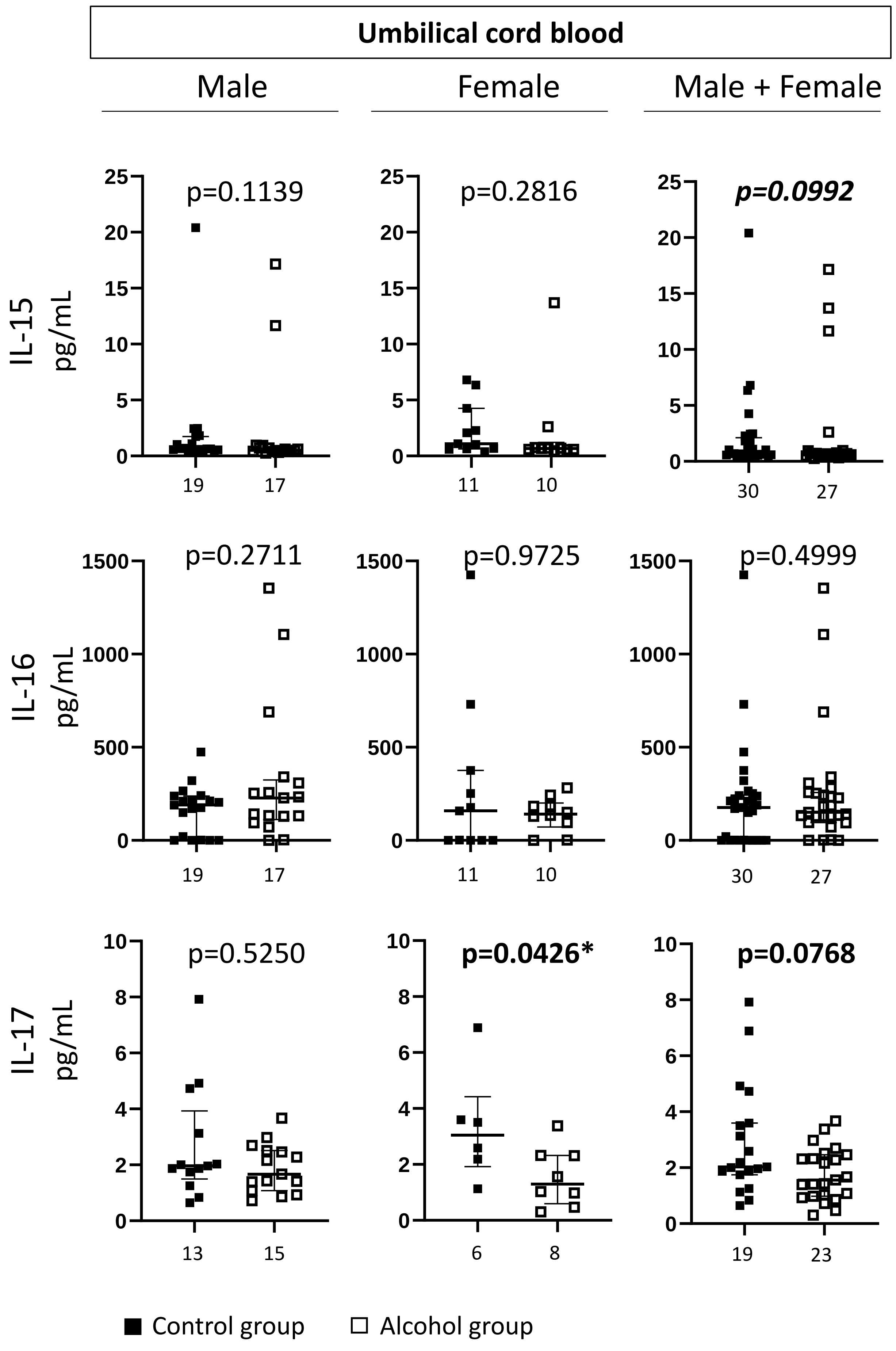

#### Chemokines/cytokines

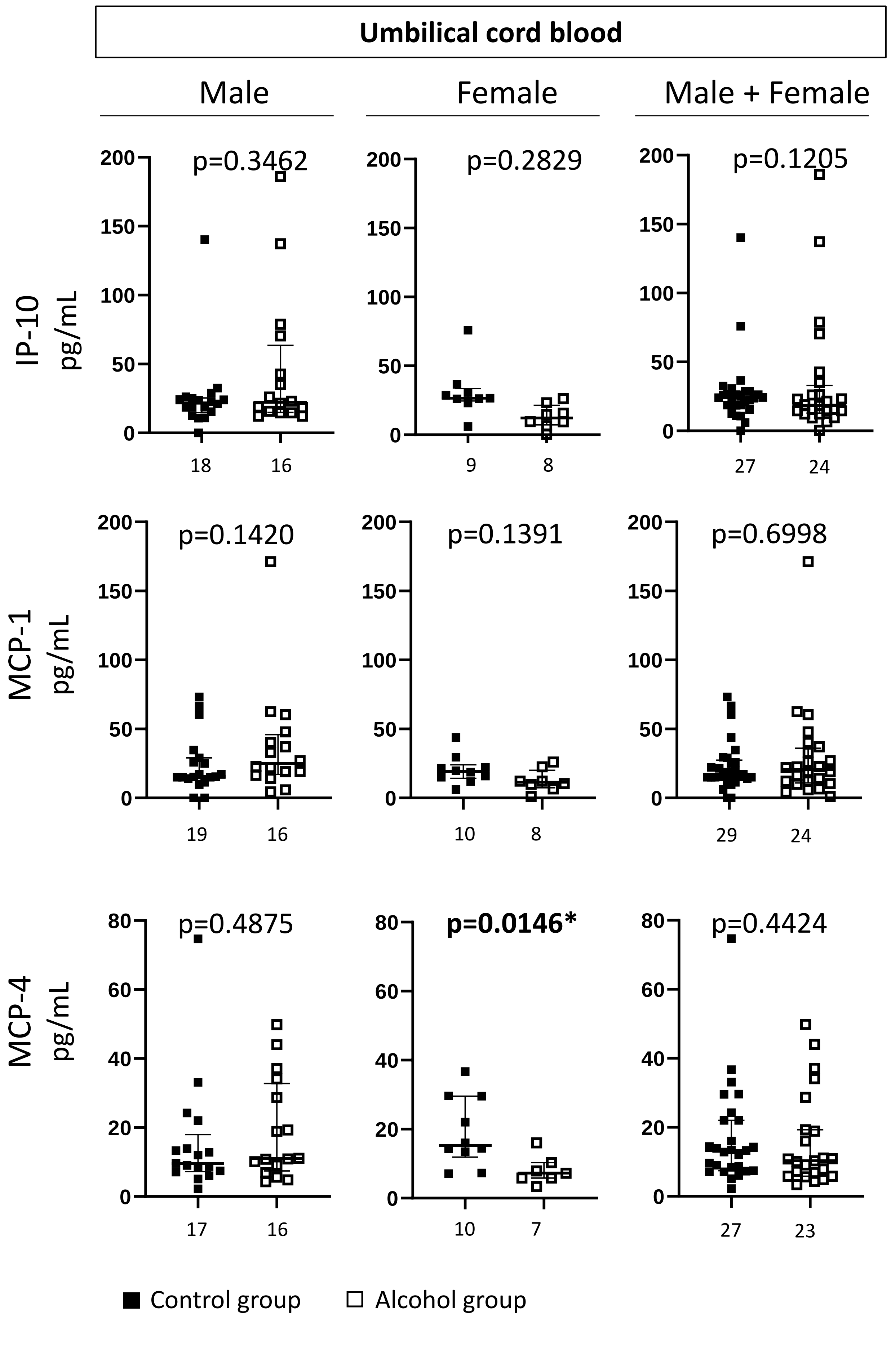

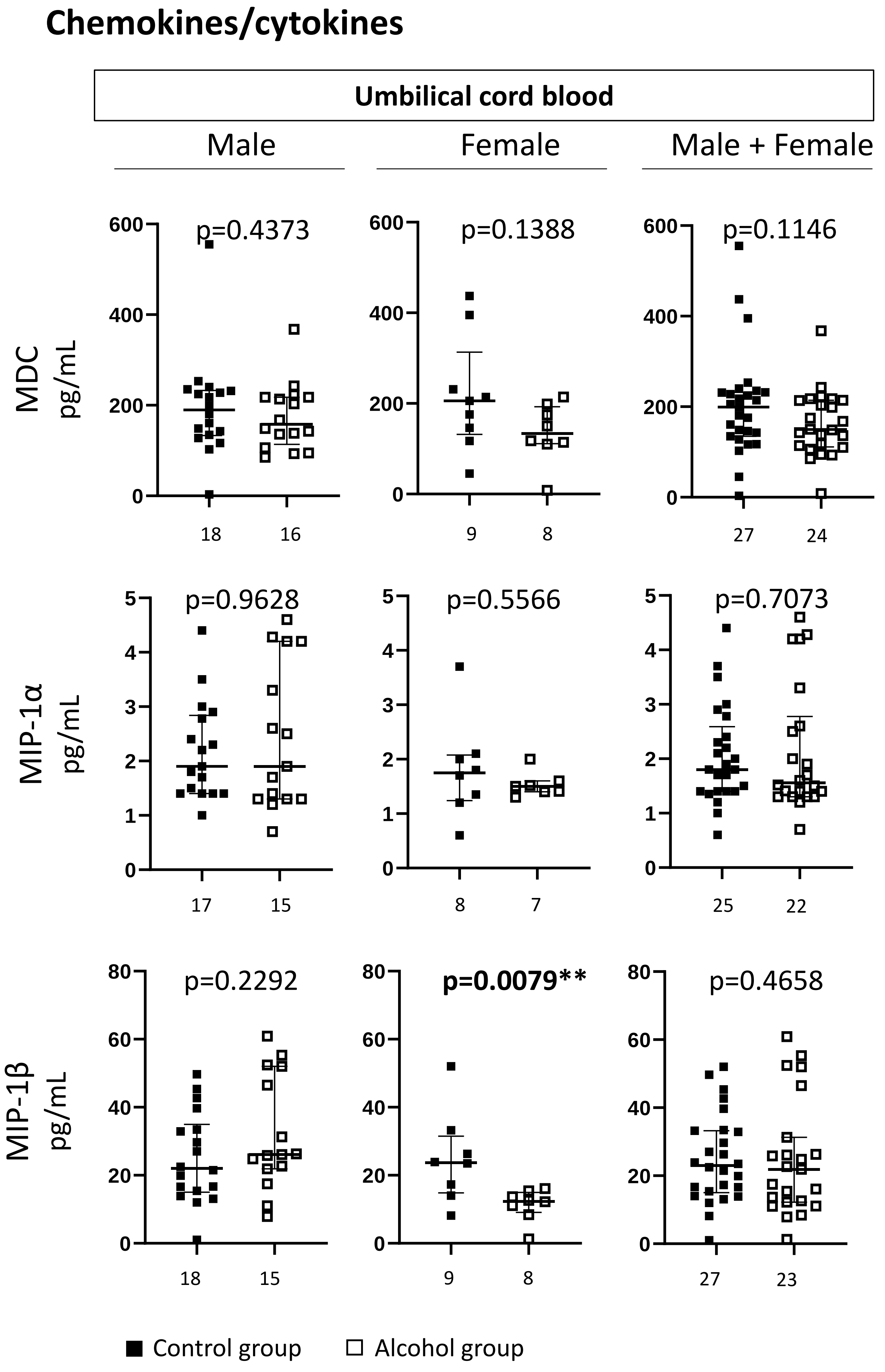
Chemokines/cytokines
Umbilical cord blood
Male
Female
Male + Female
MDC pg/mL
p=0.4373
p=0.1388
p=0.1146
18
16
9
8
27
24
MIP-1α pg/mL
p=0.9628
p=0.5566
p=0.7073
17
15
8
7
25
22
MIP-1β pg/mL
p=0.2292
p=0.0079**
p=0.4658
18
15
9
8
27
23
Control group
Alcohol group

#### Chemokines/cytokines

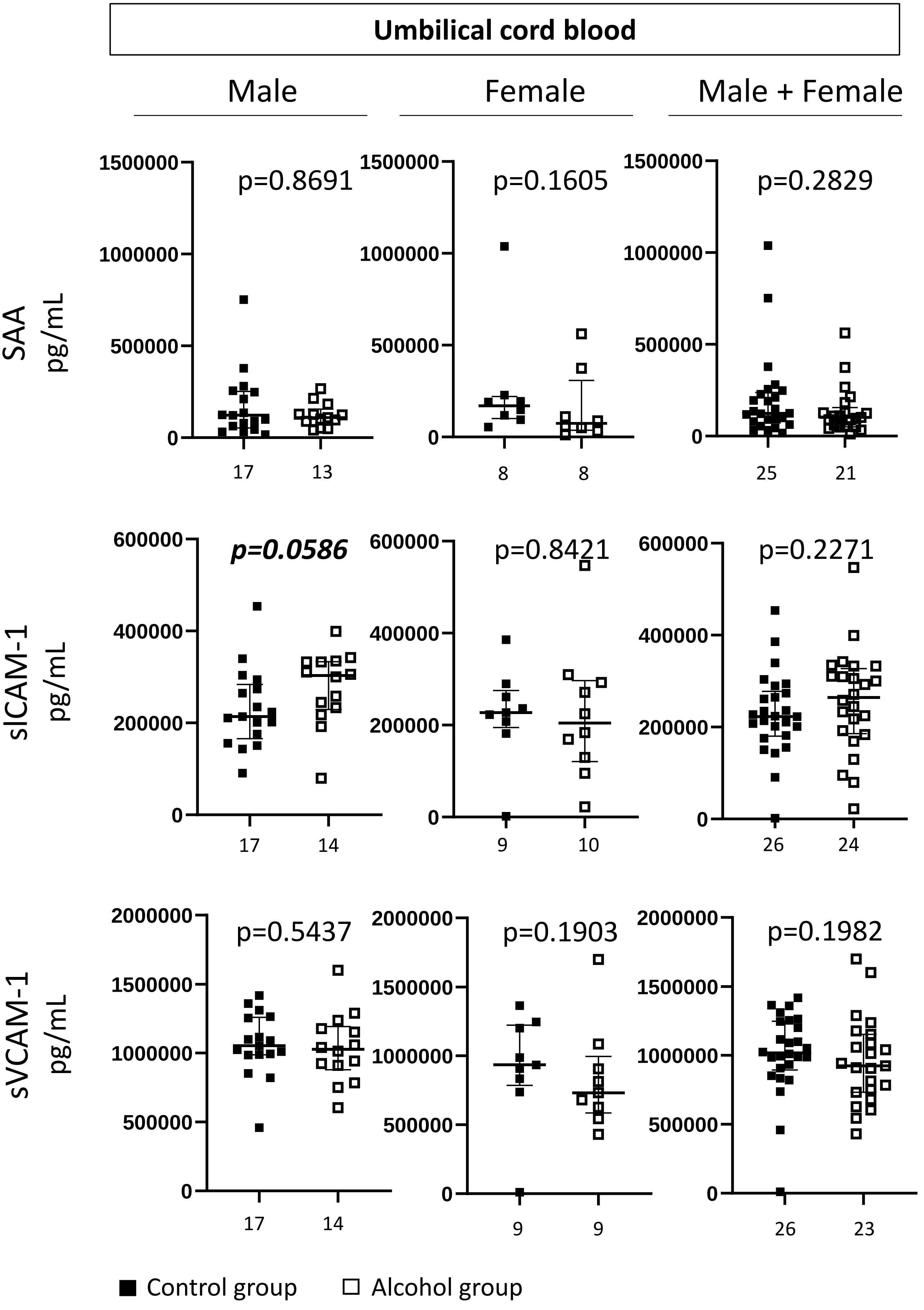

### Chemokines/cytokines

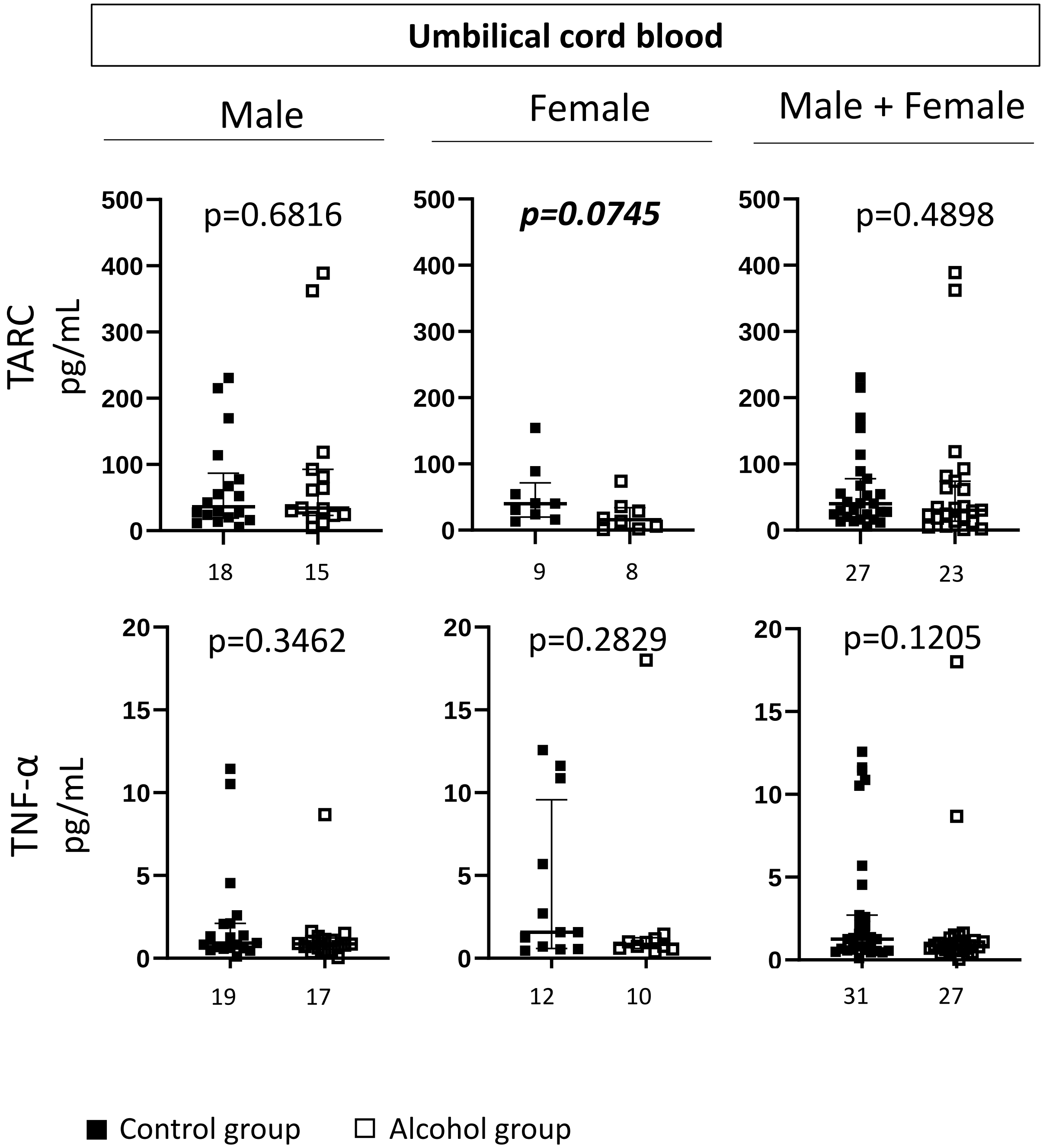


**Supplementary Figure 2. Levels of angiogenic factor, cytokine and chemokine in umbilical cord blood from Control and IUAE groups.** Numbers in abscissa indicate the number of values (n) per group. Different n values between two factors for a given group, result from missing data. Values from Control and IUAE groups were compared using the Mann-Whitney test. p<0.05 was considered statistically significant (*) and 0.05<p<0.1 considered as a trend.
